# Comparison of the immunogenicity of BNT162b2 and CoronaVac COVID-19 Vaccines in Hong Kong

**DOI:** 10.1101/2021.10.28.21265635

**Authors:** Chris Ka Pun Mok, Carolyn A Cohen, Samuel M.S. Cheng, Chunke Chen, Kin-On Kwok, Karen Yiu, Tat-On Chan, Maireid Bull, Kwun Cheung Ling, Zixi Dai, Susanna S Ng, Grace Chung-Yan Lui, Chao Wu, Gaya K. Amerasinghe, Daisy W Leung, Samuel Yeung Shan Wong, Sophie A Valkenburg, Malik Peiris, David S Hui

## Abstract

**Background:** Few head-to-head evaluations of immune responses to difference vaccines have been reported.

**Methods:** Surrogate virus neutralization test (sVNT) antibody levels of adults receiving either 2 doses of BNT162b2 (n=366) or CoronaVac (n=360) vaccines in Hong Kong were determined. An age-matched subgroup (BNT162b2 (n=49) vs CoronaVac (n=49)) were tested for plaque reduction neutralizing (PRNT) and spike binding antibody and T cell reactivity in peripheral blood mononuclear cells (PBMC).

**Findings:** One month after the second dose of vaccine, BNT162b2 elicited significantly higher PRNT_50_, PRNT_90_, sVNT, spike receptor binding, spike N terminal domain binding, spike S2 domain binding, spike FcR binding and antibody avidity levels than CoronaVac. The geometric mean PRNT_50_ titres in those vaccinated with BNT162b2 and CoronaVac vaccines were 251.6 and 69.45 while PRNT_90_ titres were 98.91 and 16.57, respectively. All of those vaccinated with BNT162b2 and 45 (91.8%) of 49 vaccinated with CoronaVac achieved the 50% protection threshold for PRNT_90._ Allowing for an expected seven-fold waning of antibody titres over six months for those receiving CoronaVac, only 16.3% would meet the 50% protection threshold versus 79.6% of BNT162b2 vaccinees. Age was negatively correlated with PRNT_90_ antibody titres. Both vaccines induced SARS-CoV-2 specific CD4^+^ and CD8^+^ T cell responses at 1-month post-vaccination but CoronaVac elicited significantly higher structural protein-specific CD4^+^ and CD8^+^ T cell responses.

**Conclusion:** Vaccination with BNT162b2 induces stronger humoral responses than CoronaVac. CoronaVac induce higher CD4^+^ and CD8^+^ T cell responses to the structural protein than BNT162b2.

**Summary At a Glance:** Through the head-to-head comparison, vaccination with BNT162b2 induces significantly higher levels of SARS-CoV-2 specific binding and neutralizing antibody responses when compared to CoronaVac. CoronaVac induce higher CD4^+^ and CD8^+^ T cell responses to the structural protein than BNT162b2.

## INTRODUCTION

SARS-CoV-2, the cause of COVID-19, emerged in late 2019 leading to a devastating pandemic (1,2). Over 240 million COVID-19 infections including 4.9 million deaths have been reported to the World Health Organization as of October 25^th^, 2021 (3). Many COVID-19 vaccines were rapidly developed, evaluated and deployed with over 4 billion doses of COVID-19 vaccines administered worldwide so far (3). These include inactivated whole virus, lipid nanoparticle (LNP)-encapsulated mRNA, adenovirus-vectored and protein sub-unit vaccines. Virus neutralizing antibodies correlate with protection from re-infection following natural infection as well as vaccination (4, 5) and in experimental animal models (6). T cell immune responses are consistently elicited after natural infection and correlate with reduced disease severity in humans and reduced viral loads in non-human primates (7-11).

The safety, immunogenicity and efficacy of these vaccines have been evaluated in separate clinical trials (12-14) but there are few “head-to-head” comparisons of different vaccines. Here, we compare the humoral and cellular responses from vaccinees who received either BNT162b2 or CoronaVac.

## MATERIALS AND METHODS

### Cohort study design and participants

Healthy adults aged between 18-79 years were recruited in Hong Kong SAR, China at vaccination centers at the Chinese University of Hong Kong Medical Centre, Prince of Wales Hospital and Kowloon Bay Vaccination Station between March10 and August 31, 2021. Those with previous COVID-19 infection were excluded. The participants received two doses of either BNT162b2 [Comirnaty(BioNTech)] vaccine (21 day interval between the two doses) or CoronaVac (Sinovac) vaccine (28 day interval between the two doses), according to the manufacturer’s recommendation. Demographic information was collected from each participant prior to vaccination (Supplementary Table 1). Ten ml of heparinized blood were collected from each donor before vaccination and at one month after receiving the second vaccine dose.

### Immunological Assays

Humoral and cellular immune responses were examined to compare the levels of SARS-CoV-2 neutralizing, binding and FcR^+^ binding antibody in plasma and T cell reactivity in peripheral blood mononuclear cells (PBMC) between the two vaccine groups. Details of specimen storage and processing and the serological methods used in ELISA for spike receptor binding domain (RBD), spike N terminal domain (NTD) binding, S2 and N protein antibodies, ELISA for FcγRIIIa-binding and antibody avidity, surrogate virus neutralization test (sVNT), plaque reduction neutralization test (PRNT), and SARS-CoV-2-specific T cells by Intracellular Cytokine Staining (ICS) are provided in the supplemental file.

### Statistical analysis

Methods used for statistical analysis are provided in the supplemental file.

### Role of the funding source

The sponsor of the study had no role in study design, data collection, data analysis, data interpretation, or writing of the report. The corresponding authors had full access to all the data in the study and had final responsibility for the decision to submit for publication.

## RESULTS

Adult volunteers (age range 18-79 years) were recruited between March 10 and August 31, 2021; 366 of whom received two doses of BNT162b2 and 360 received two doses of CoronaVac. The mean (SD) age of the two groups were 45.01 (13.16) and 51.78 (9.92) years respectively (p<0.0001). Demographic information is summarized in Supplementary Table 1. We used sVNT, which has high overall concordance to the PRNT (15,16), to examine the level of SARS-CoV-2 specific neutralizing antibody from the plasma samples collected before and at 1 month after the 2nd dose of vaccination. The plasma samples from all vaccinees before vaccination were negative in sVNT (data not shown). The mean % inhibition in the sVNT test in post-vaccine plasma for BNT1626 and CoronaVac was 93.63% (standard deviation (SD) 7.92) and 52.11% (SD 22.06), respectively (p<0.0001) (Figure 1A).

**Figure 1.**
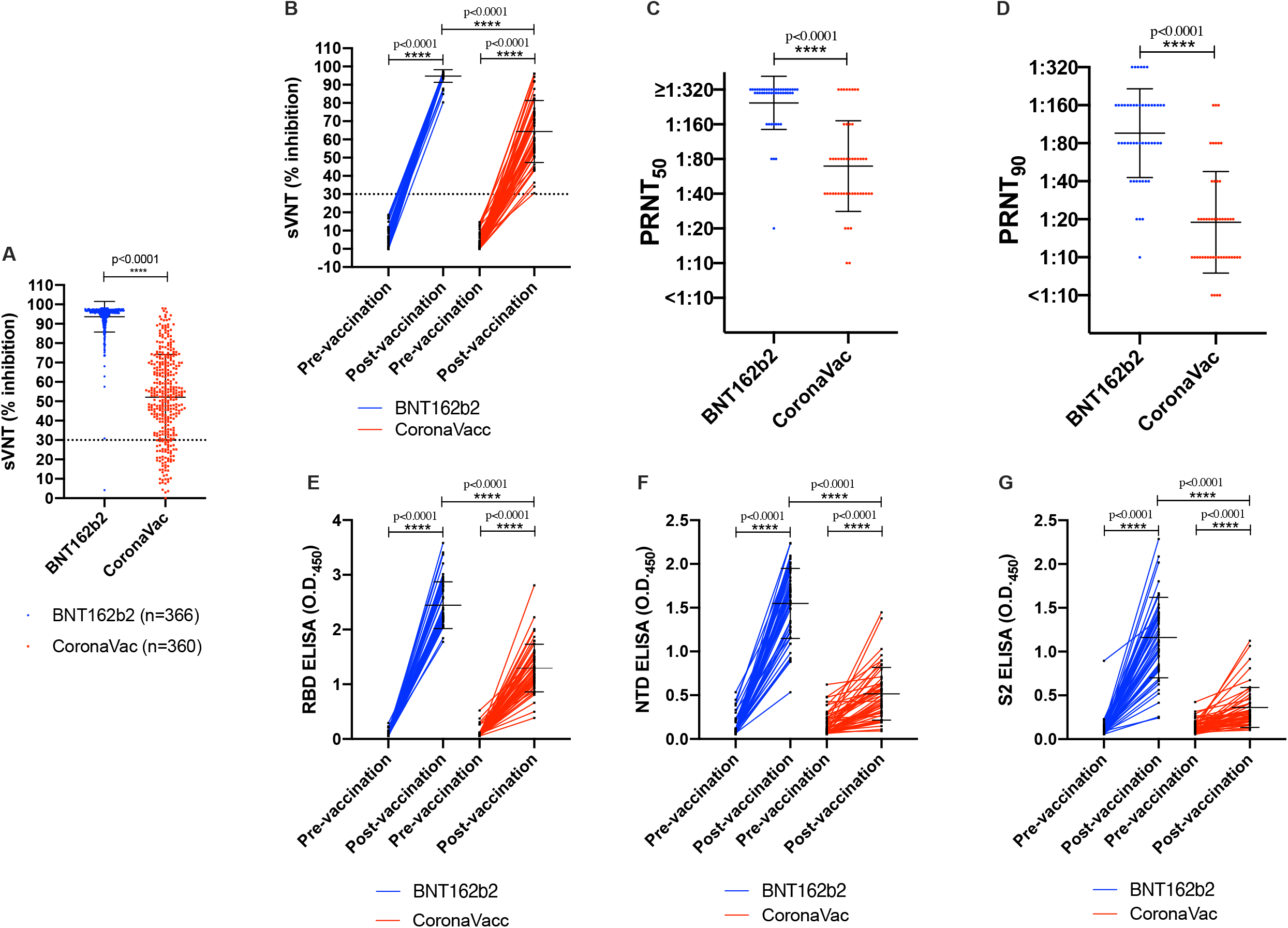
Antibody responses of individuals before and after BNT162b2 or CoronaVac vaccination. The percentage of inhibition were detected by surrogate virus neutralization test (sVNT) from the plasma collected from adult individuals who received two doses of BNT162b2 (n=366) or received two doses of CoronaVac (n=360) (A). Various antibody responses were further determined from the plasma from an age-matched subgroup of (A) (49 vs 49). (B) The percentage of inhibition from the plasma of pre-vaccination and 1 month after two doses of vaccination were tested by sVNT. The dash line at 30% indicates the negative threshold of the sVNT. Comparison of the (C) PRNT_50_ and (D) PRNT_90_ from the plasma collected at 1 month after two doses of vaccination between the BNT162b2 and CoronaVac groups. The levels of (E) RBD-specific (F) NTD-specific and (G) S2-specific IgG antibodies from the plasma of pre-vaccination and 1 month after two doses of vaccination were tested by ELISAs. **** indicates p<0.0001

From the data above, we estimated that a sample size of 49 for each vaccine group provided statistical power higher than 80% with 95% confidence level to discern differences between the two vaccines. To avoid age-related bias, we randomly 49 selected age-matched samples from BNT162b2 or CoronaVac vaccinees for more detailed analysis. The demographic, underlying co-morbidities and other potential risk-factors were shown in Table 1. The mean % inhibition in the sVNT test in post-vaccine plasma for BNT1626 and CoronaVac was 94.8% (standard deviation (SD) 3.45) and 63.9% (SD 16.72), respectively (p=7.71×10^−17^) (Figure 1B).

**Table 1.**
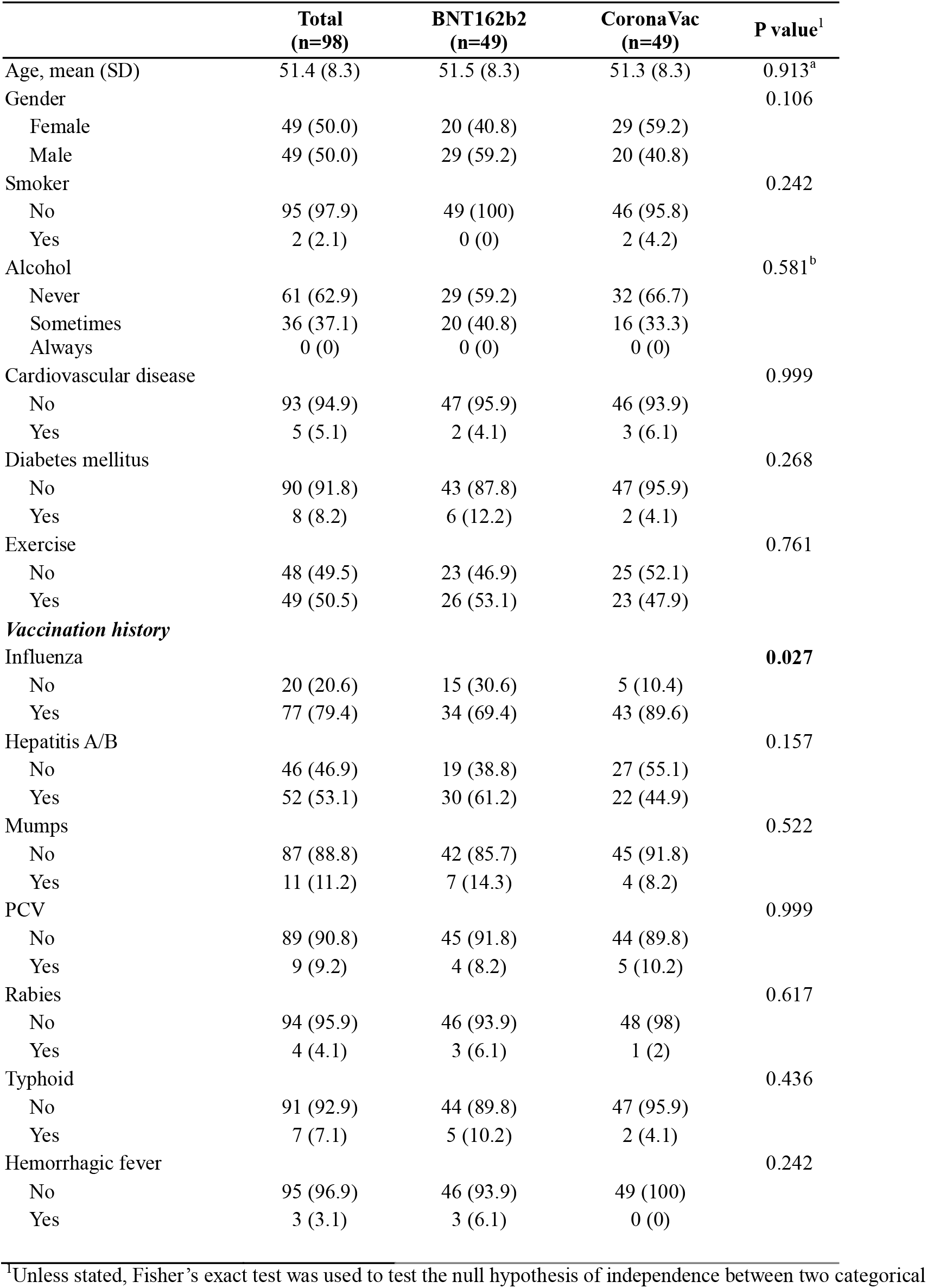
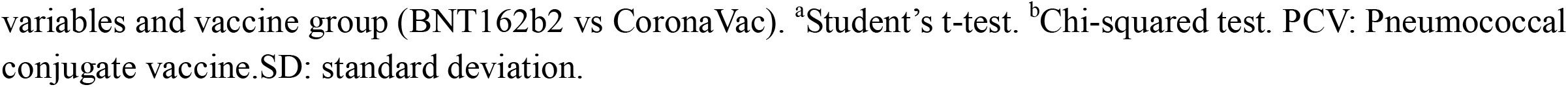
Comparison of characteristics between the two vaccine groups (n=98).

|  | **Total**<br>**(n=98)** | **BNT162b2**<br>**(n=49)** | **CoronaVac**<br>**(n=49)** | **P value<sup>1</sup>** |
| --- | --- | --- | --- | --- |
| Age, mean (SD) | 51.4 (8.3) | 51.5 (8.3) | 51.3 (8.3) | 0.913<sup>a</sup> |
| Gender |  |  |  | 0.106 |
| Female | 49 (50.0) | 20 (40.8) | 29 (59.2) |  |
| Male | 49 (50.0) | 29 (59.2) | 20 (40.8) |  |
| Smoker |  |  |  | 0.242 |
| No | 95 (97.9) | 49 (100) | 46 (95.8) |  |
| Yes | 2 (2.1) | 0 (0) | 2 (4.2) |  |
| Alcohol |  |  |  | 0.581<sup>b</sup> |
| Never | 61 (62.9) | 29 (59.2) | 32 (66.7) |  |
| Sometimes | 36 (37.1) | 20 (40.8) | 16 (33.3) |  |
| Always | 0 (0) | 0 (0) | 0 (0) |  |
| Cardiovascular disease |  |  |  | 0.999 |
| No | 93 (94.9) | 47 (95.9) | 46 (93.9) |  |
| Yes | 5 (5.1) | 2 (4.1) | 3 (6.1) |  |
| Diabetes mellitus |  |  |  | 0.268 |
| No | 90 (91.8) | 43 (87.8) | 47 (95.9) |  |
| Yes | 8 (8.2) | 6 (12.2) | 2 (4.1) |  |
| Exercise |  |  |  | 0.761 |
| No | 48 (49.5) | 23 (46.9) | 25 (52.1) |  |
| Yes | 49 (50.5) | 26 (53.1) | 23 (47.9) |  |
| ***Vaccination history*** |  |  |  |  |
| Influenza |  |  |  | **0.027** |
| No | 20 (20.6) | 15 (30.6) | 5 (10.4) |  |
| Yes | 77 (79.4) | 34 (69.4) | 43 (89.6) |  |
| Hepatitis A/B |  |  |  | 0.157 |
| No | 46 (46.9) | 19 (38.8) | 27 (55.1) |  |
| Yes | 52 (53.1) | 30 (61.2) | 22 (44.9) |  |
| Mumps |  |  |  | 0.522 |
| No | 87 (88.8) | 42 (85.7) | 45 (91.8) |  |
| Yes | 11 (11.2) | 7 (14.3) | 4 (8.2) |  |
| PCV |  |  |  | 0.999 |
| No | 89 (90.8) | 45 (91.8) | 44 (89.8) |  |
| Yes | 9 (9.2) | 4 (8.2) | 5 (10.2) |  |
| Rabies |  |  |  | 0.617 |
| No | 94 (95.9) | 46 (93.9) | 48 (98) |  |
| Yes | 4 (4.1) | 3 (6.1) | 1 (2) |  |
| Typhoid |  |  |  | 0.436 |
| No | 91 (92.9) | 44 (89.8) | 47 (95.9) |  |
| Yes | 7 (7.1) | 5 (10.2) | 2 (4.1) |  |
| Hemorrhagic fever |  |  |  | 0.242 |
| No | 95 (96.9) | 46 (93.9) | 49 (100) |  |
| Yes | 3 (3.1) | 3 (6.1) | 0 (0) |  |

<sup>1</sup>Unless stated, Fisher's exact test was used to test the null hypothesis of independence between two categorical
variables and vaccine group (BNT162b2 vs CoronaVac). <sup>a</sup>Student's t-test. <sup>b</sup>Chi-squared test. PCV: Pneumococcal conjugate vaccine. SD: standard deviation.

PRNT is the gold-standard method to evaluate virus neutralization. All vaccinees developed detectable PRNT_50_ antibody titers. BNT162b2 vaccinees had significantly higher geometric mean titers (GMT) of 251.60 (±1SD range from 147 to 432) compared to 69.45 (±1SD range from 28 to 172) for the CoronaVac group (p=1.24×10^−9^) (Figure 1C). The corresponding PRNT_90_ antibody responses for BNT162b2 (GMT 98.91, ±1SD range from 44 to 221) was significantly higher than for CoronaVac vaccines (GMT 16.57, ±1SD range from 5 to 54) (p=5.82×10^−10^) (Figure 1D). It has been suggested that the neutralizing antibody titer associated with protection from re-infection in 50% of individuals is 20% of the mean convalescent antibody levels (5). Using RT-PCR confirmed convalescent symptomatic patients sera (16), using the same PRNT methods, we estimated that this 20% convalescent antibody titer threshold for 50% protection from re-infection for PRNT_90_ was 1:8.75 (95% CI 1:6.6-1:11.6) (Supplementary Figure 1). Thus, we estimated that all 49 vaccinated with BNT162b2 and 45 (91.8%) of 49 of those vaccinated CoronaVac achieved the 50% protection threshold, one month after the second dose of vaccination. Neutralizing antibody levels reportedly fell by 7.3-fold within 6 months of CoronaVac immunization (17). Thus we estimate that only 8 (16.3%) of 49 CoronVac vaccines would retain protective levels of neutralizing antibody 6 months post vaccination while 79.6% of BNT162b2 vaccinees would do so, assuming comparable waning of antibody in BNT162b2 vaccinees.

Adjusting for age, gender and history of other vaccine uptake, subjects who received CoronaVac had a significantly lower PRNT_90_ value compared to those who received BNT162b2 (Supplementary Table 2). Stratifying regression analysis by choice of vaccine, older age was independently associated with lower PRNT_90_ values with either vaccine (Supplementary Table 3).

Antibodies against different domains of the spike can facilitate protection (18, 19, 20). Thus, we further tested the levels of IgG antibodies specifically binding to the RBD, NTD and S2 domains of the spike protein, respectively. While both vaccines elicited significant increases in these antibodies as detected by ELISA, BNT162b2 vaccine elicited significantly higher antibody levels to all three antigens (Figure 1E-G). As expected from a whole virus vaccine which also contains the virus N protein, 59.2% (29/49) and 40.8% (20/49) of post-vaccination plasma from CoronaVac group were positive in ELISA against the full length (aa 44-419) and C-terminal domain of the N protein (Supplementary Figure 2). One plasma sample from a BNT162b2 vaccinee had high ELISA binding antibody to N protein (but not C-terminal domain of N) in post-vaccination plasma, but was negative for other viral proteins (ORF8, data not shown), and therefore may represent cross-reactivity in the N antibody assay, perhaps with other seasonal coronaviruses.

Antibodies which bind to Fc Receptors on effector cells such as natural killer (NK) cells may stimulate antibody dependent cell cytotoxicity (ADCC) upon FcgRIIIa binding of virus-antibody complexes leading to down-stream signaling. Antigen specific FcgRIIIa binding significantly corelates with NK cell function of degranulation, killing and cytokine production (21). Furthermore, increased ADCC function is associated with mRNA (22) and protein (23) vaccination and reduced morbidity during infection (24, 25), and is therefore regarded as one of the indicators of immune protection. We found that recipients from both BNT162b2 or CoronaVac elicit increases of FcγRIIIa binding spike antibodies after two doses of vaccination (Figure 2A). However, the antibody level from the BNT162b2 group was significantly higher than the CoronaVac group. CoronaVac recipients had higher levels of FcγRIIIa binding antibodies to N than the BNT162b2 group after vaccination (p <0.0001) as expected, but CoronaVac N-FcγRIIIa responses were still significantly lower than COVID-19 cases (Figure 2B). FcγRIIIa binding is more likely to result in downstream signaling leading to effector function if antibodies have high avidity (26). S-specific FcγRIIIa as well as S-IgG avidity was significantly lower in CoronaVac recipients than BNT162b2 recipients or COVID-19 convalescent patients (Figure 2C, D). The average avidity of N specific FcγRIIIa from CoronaVac recipients were comparable to the patients with COVID-19 (Figure 2E).

**Figure 2.**
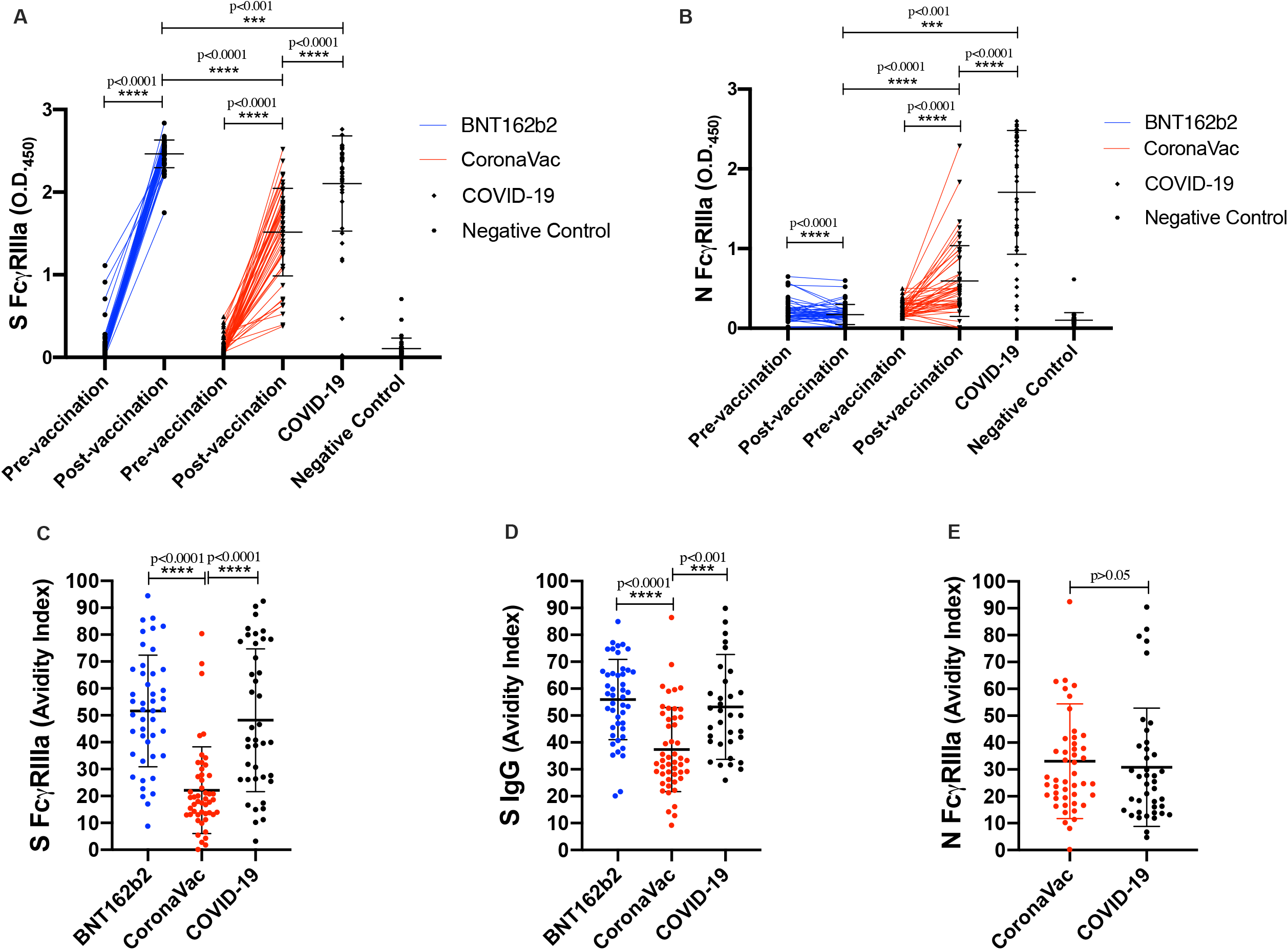
FcγRIIIa binding antibodies and IgG avidity in the BNT162b2 and CoronaVac groups. The levels of FcγRIIIa binding antibodies and their avidity were detected from the plasma collected from adult individuals who received two doses of BNT162b2 (n=49) or received two doses of CoronaVac (n=49). Recovered COVID-19 cases (n=34, timepoint 56 days post infection +/- 17 days (mean+/- SD)), and healthy adults negative for SARS-CoV-2 (n=40) were served as positive and negative control respectively. The levels of (A) FcγRIIIa binding S antibodies and (B) FcγRIIIa binding N antibodies were tested from the plasma collected before and at 1 month after 2 doses of vaccination. The avidity indexes of (A) S FcγRIIIa, (B) N FcγRIIIa and (C) S IgG were determined as the proportion of antibodies remaining after 3x washes with 8M urea, compared to the total FcγRIIIa-binding antibodies to each protein. **** indicates p<0.0001

We collected PBMCs from the same subgroup where possible, making a cohort of 25 vaccinees who received two doses of BNT162b2 and 30 with two doses of CoronaVac. Overlapping peptides of the structural proteins including S, N, Envelope and Matrix (SNEM) or S only pools were used to induce specific T cell responses in the PBMCs. The structural and S specific CD4^+^ and CD8^+^ T cell responses were quantified by flow cytometry (Supplementary Figure 3) from paired samples at pre and post vaccination of CoronaVac and BNT162b2 while RT-PCR confirmed COVID-19 patients were served as positive controls. The average magnitude of the post vaccination CD4^+^ and CD8^+^ T cell responses after stimulation of SNEM peptides was significantly higher in CoronaVac group compared those received BNT162b2 (Figure 3A and B). The magnitude of the post vaccination CD4^+^ T cell response after stimulation of S only peptides was also significantly higher in CoronaVac group but their CD8^+^ T cell responses was comparable to the BNT162b2 group (Supplementary Figure 4 A and B). Overall, the proportion of subjects that had detectable post vaccination T cell responses (% of IFNγ^+^ cell is higher than 0.001), termed ‘responders’ was higher in CoronaVac recipients than BNT162b2 recipients, for either SNEM peptides or S peptides alone (Table 2).

**Table 2.** Structural and spike-specific T cell responses assessed by intracellular staining for interferon γ.

| % (n) |  | Pre vaccination |  |  | Post vaccination |  |  |  |
| --- | --- | --- | --- | --- | --- | --- | --- | --- |
| **Structural protein** | | Responder | Non-responder | % IFN$\gamma^+$ of T cells | Responder | Non-responder | % IFN$\gamma^+$ of T cells | % IFN$\gamma^+$ of T cells<br>Fold change |
| CD4$^+$ T cells | BNT162b2 | 28% (7) | 72% (18) | 0.02032 +/- 0.04548 | 32% (8) | 68% (17) | 0.0288 +/- 0.054 | 13.40 +/- 27.59 |
|  | CoronaVac | 50% (15) | 50% (15) | 0.0431 +/- 0.06474 | 83.3% (25) | 16.7% (5) | 0.11217 +/- 0.10786 | 72.39 +/- 112.04 |
| CD8$^+$ T cells | BNT162b2 | 20% (5) | 80% (20) | 0.03081 +/- 0.10046 | 32% (8) | 68% (17) | 0.03246 +/- 0.07995 | 21.05 +/- 60.39 |
|  | CoronaVac | 33.3% (10) | 66.7% (20) | 0.0189 +/- 0.03491 | 63.3% (19) | 36.7% (11) | 0.1104 +/- 0.1167 | 77.20 +/- 103.89 |
| **Spike protein** | | Responder | Non-responder | % IFN$\gamma^+$ of T cells | Responder | Non-responder | % IFN$\gamma^+$ of T cells | % IFN$\gamma^+$ of T cells<br>Fold change |
| CD4$^+$ T cells | BNT162b2 | 52% (13) | 48% (12) | 0.05808 +/- 0.1085 | 44% (11) | 56% (14) | 0.04656 +/- 0.07888 | 25.01 +/- 51.82 |
|  | CoronaVac | 60% (18) | 40% (12) | 0.0597 +/- 0.08131 | 73.3% (22) | 26.7% (8) | 0.0875 +/- 0.08939 | 37.65 +/- 64.64 |
| CD8$^+$ T cells | BNT162b2 | 36% (9) | 64% (16) | 0.04053 +/- 0.0729 | 36% (9) | 64% (16) | 0.09492 +/- 0.19278 | 39.83 +/- 80.84 |
|  | CoronaVac | 46.7% (14) | 53.3% (16) | 0.0286 +/- 0.07923 | 50% (15) | 50% (15) | 0.0744 +/- 0.10961 | 47.94 +/- 87.52 |

**Figure 3.**
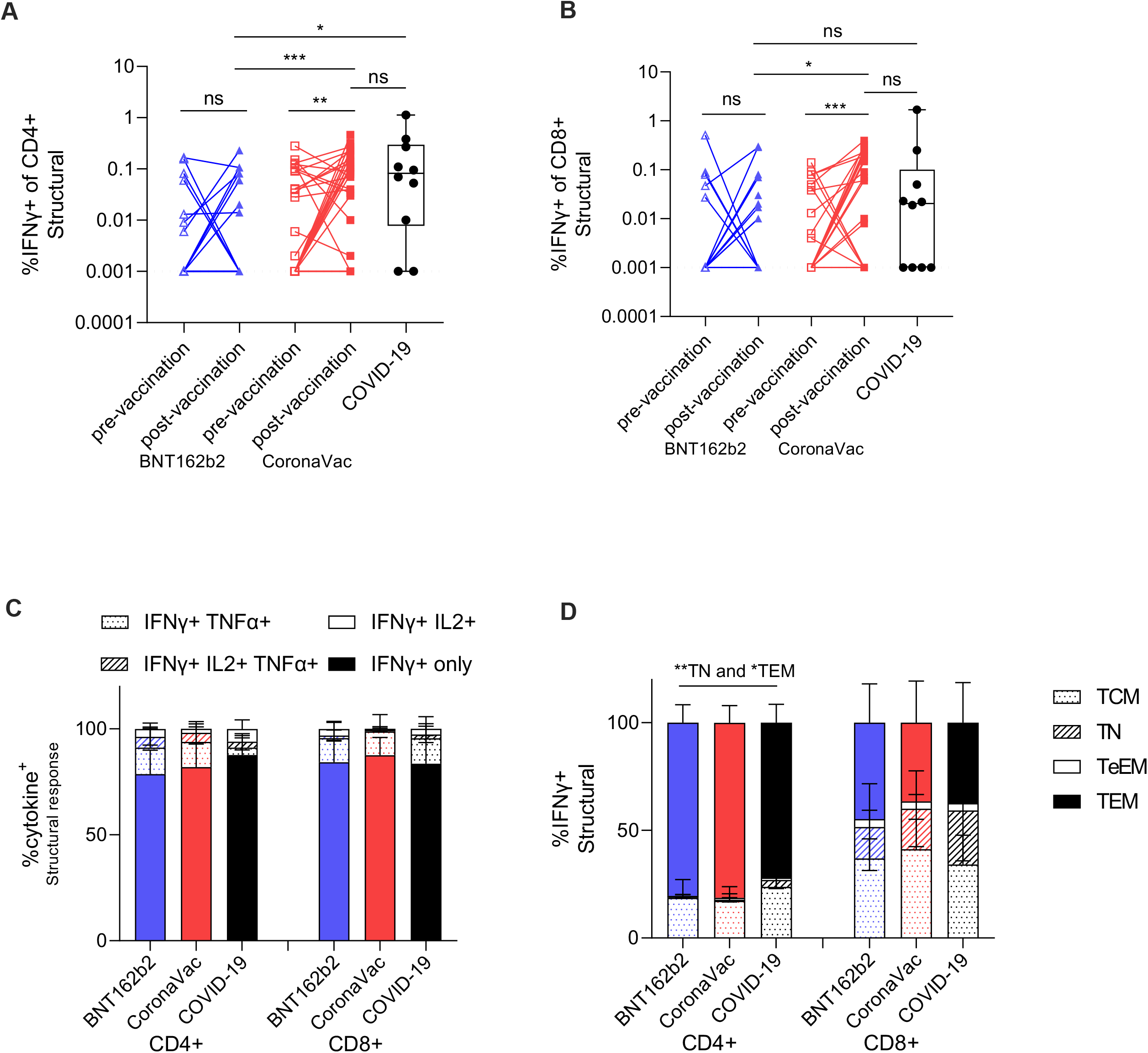
T cell responses post vaccination are comparable between BNT162b2 or CoronaVac. PBMCs from pre- (day 0) and post-vaccination (day 30 post after second dose) of BNT162b2 mRNA (pre-vaccine n=25, post-vaccine n=25) and CoronaVac (pre-vaccine n=30, post-vaccine n=30) and recovered COVID-19 cases (n=10, timepoint 59 days post infection +/- 20 days (mean+/- SD)) were stimulated with pooled structural (SNEM) peptides or a DMSO control. The percentage of (A) IFNγ^+^CD4^+^ and (B) IFNγ^+^CD8^+^ T cell were measured by flow cytometry. Dotted lines represent limit of detection following DMSO background subtraction (IFNγ of CD4^+^=0.001, IFNγ of CD8^+^=0.001. (C) The proportion of IFNγ producing IL2 and TNFα CD4^+^ and CD8^+^ T cells post vaccination. (D) The phenotype (by CCR7 and CD45RA) of IFNγ responses for T effector memory (TEM), central memory (TCM), terminal effector memory (TeEM) or naïve (TN) CD4^+^ and CD8^+^ T cells post vaccination. Bars represent the mean values, and error bars represent SD. Statistical significance was determined by paired t-test between pre-versus post-vaccination timepoint samples, and Kruskal-Wallis test for multiple comparisons between vaccines, and COVID-19 patients. *p<0.05, **<0.01, ***<0.001.

Post vaccination polyfunctional cytokine production, including TNFα and IL-2 by T cells in the vaccine responders was equivalent between the two vaccine types and convalescent COVID-19 samples after stimulation of either the SNEM or S only peptides (Figure 3C; Supplementary Figure 4C). The memory phenotype of IFNγ^+^ CD4^+^ T cells in both vaccine groups showed significantly higher percentage of T effector memory (TEM) (p<0.05) and lower in T naïve (TN) (p<0.01) than the COVID-19 recovered patients (Figure 3D). Interestingly, S-specific CD4^+^ T cell responses memory phenotypes further showed TEM>TCM (T central memory) in the CoronaVac group compared to BNT126b2 which may impact recall at infection and long-term cellular memory (Supplementary Figure 4D).

## DISCUSSION

Using a cohort of RT-PCR confirmed convalescent sera from COVID-19 patients, using the assays we have used in the present study, we estimated the 20% convalescent antibody titer threshold for 50% protection from re-infection for PRNT_90_ was 1:8.75 (95% CI 1:6.6-1:11.6) (5,16). We conclude that all those vaccinated with BNT162b2 and 91.8% of those vaccinated CoronaVac achieved the 50% protection threshold one month after the second dose of vaccination. One month post-second dose likely represents the peak antibody response, beyond which antibody titers are likely to wane. It was reported that neutralizing antibody titres wane 7.3-fold within 6 months after CoronaVac vaccination (17) but comparable data is not available from BNT126b2. If we adjust for a 7.3-fold waning of antibody titres for both vaccines, we estimate that only 8 (16.3%) of 49 receiving CoronaVac vaccines meet the protective threshold while 39 (79.6%) of 49 of those receiving BNT162b2 do so, 6 months post-vaccination. This difference in immunogenicity may explain reported difference in vaccine efficacy between the two vaccines (27-30). Many Phase 3 studies assessed protection within 1 to 3 months after the second dose of vaccine and may not reflect the impact of waning immunity. Furthermore, some of the virus variants (e.g. B.1.351 Beta variant; P1 Gamma variant) circulating in parts of the world lead to an 8 to 10-fold reduction in neutralizing titers and this is likely to further compromise protection afforded by CoronaVac vaccines with its weaker immunogenicity. Our results suggest that booster doses may be required for older CoronaVac recipients and such booster doses. A third dose of CoronaVac vaccine appears to boost antibody levels (17) and our data suggest that these may be needed for older CoronaVac recipients.

The average magnitude of post vaccination responses was higher in CoronaVac subjects for structural and S-specific T cell responses. Since it is likely that T cell responses are important in limiting severity and fatal outcomes (7-11), both vaccines may be effective in preventing such adverse outcomes of COVID-19. This is consistent with high levels of protection against hospitalization and death in CoronaVac vaccines observed in Chile and Turkey despite lower antibody neutralization titers than BNT162b2 (30, 31). In animal models of re-infection, spike-specific CD8^+^ T cell responses can compensate for inadequate antibody responses (24) and are also highly cross-reactive to different variants of concern (32). Therefore, SARS-CoV-2 specific T cells may provide an additional contribution to the immune correlate of protection. An advantage of inactivated vaccines is that they also contain additional viral antigens, such as the highly abundant and immunogenic N protein which may also elicit T-cell immunity (33) and contributed to the higher responses in CoronaVac subjects here. Sampling at earlier timepoints (day 7-14) post vaccination may reveal differences in response magnitude given the phenotypic expansion of S-specific TEM CD4^+^ T cells by the BNT162b2 vaccine which may also have greater recall potential at infection (34). Longitudinal follow-up is thus necessary to confirm the long-term T cell responses between the two vaccines.

Proline mutations used in BNT162b2 vaccine to stabilize the spike protein in its pre-fusion state (35) and the serial passages in Vero cells and inactivation procedures used in the CoronaVac vaccine (36) are differences that may contribute to the differences in neutralizing antibody responses elicited by the two vaccines.

Preliminary results from another study has recently reported a marked difference of immunogenicity between BNT162b2 and CoronaVac in healthcare workers (37). However, the median age of the two groups were 37 and 47 years old respectively in that study while our subgroup was designed to be age matched. Moreover, the ADCC and T cell responses were not examined in their cohort.

There were some limitations in our study. The choice of vaccine was not randomized and there might be a selection bias in those opting for each vaccine. Our study only focused on investigating the immunogenicity at 1 month after two doses of vaccination. The durability of immune responses needs to be monitored; and indeed, this cohort will be followed up to address this question in the coming years. Our estimates to adjust for antibody waning was based on reports on CoronaVac, comparable to data for BioNTech was lacking. Thus our assumption of comparable rates of antibody waning for the two vaccines may not be correct. We did not collect plasma prior to the second dose of vaccine to assess the effect of the first dose of the vaccine, or acute phase responses, where earlier responses may account for the final post vaccine differences as our primary study endpoint was neuralization titers after vaccination. Similar comparisons between vaccines in teenagers and older adults will be needed.

## Conclusion

Our data showed that the levels of antibodies elicited by CoronaVac were significantly lower than those of BNT162b2 in PRNT_50_, PRNT_90_, sVNT, spike RBD ELISA, spike NTD ELISA and spike S2 ELISA assays, and total and high avidity FcgRIIIa spike. CoronaVac induce higher CD4^+^ and CD8^+^ T cell responses to the structural protein than BNT162b2.

## Data Availability

All data produced in the present work are contained in the manuscript

## ACKNOWLEDGMENTS

We acknowledge the technical support from Mr Huibin Lv and Mr Ho Lun Lai. We also thank Dr Fung Hong (Chinese University Medical Center), Dr Ken Tsang (Kowloon Bay community vaccination center) and Dr Beatrice Cheng (Prince of Wales Hospital) for allowing us to recruit subjects for this study. The recombinant RBD and NTD proteins were kindly provided as gifts by Prof Ian A. Wilson and Dr Meng Yuan. This project utilized an Invitrogen Attune flow cytometer funded by the Pasteur Foundation Asia.

## FIGURE LEGENDS

**Supplementary Figure 1:**
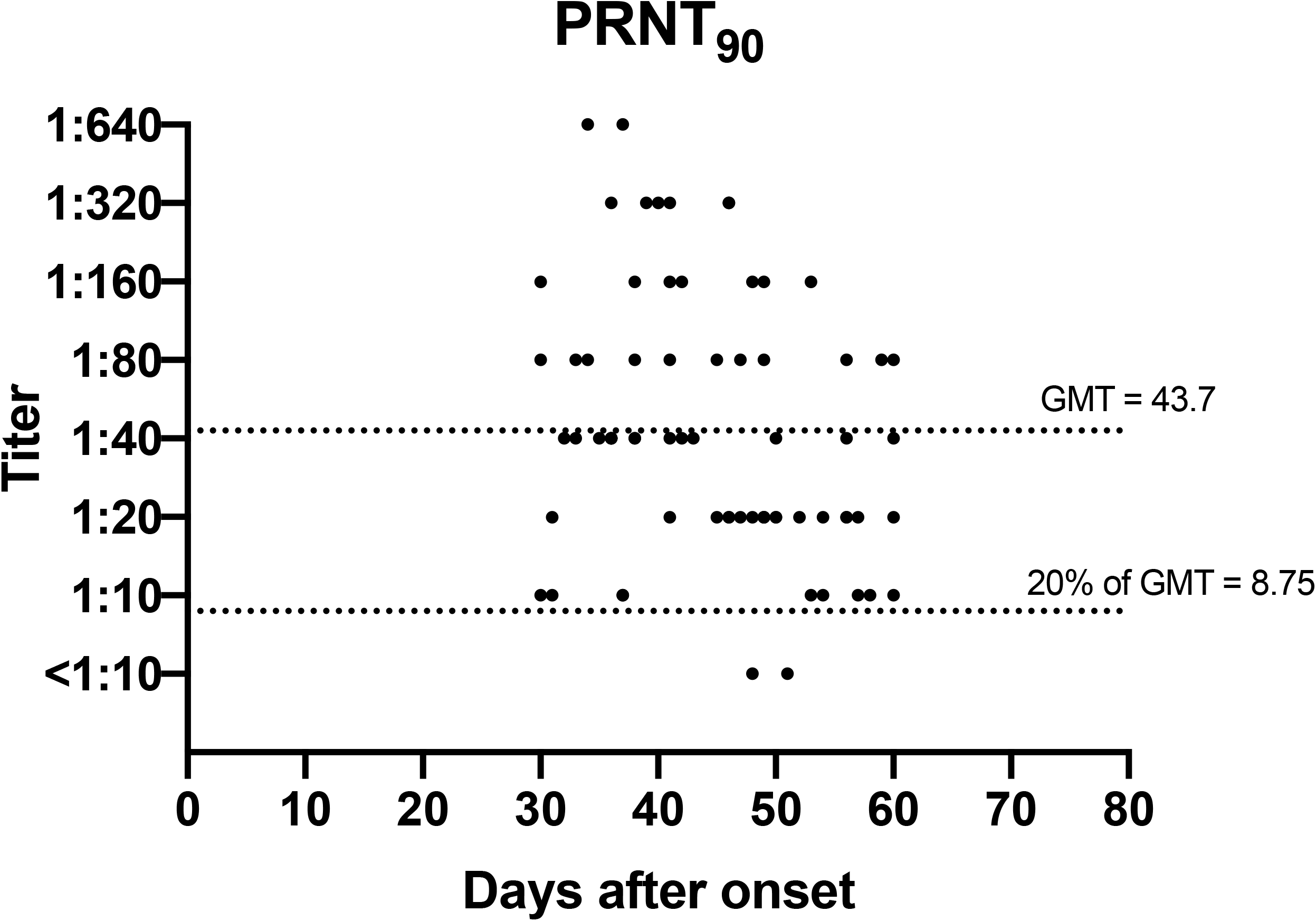
PRNT_90_ antibody titres in 70 sera from 50 symptomatic COVID-19 patients collected between 30 to 60 days after illness onset is shown. Geometric mean antibody titers (GMT) is denoted in the solid lines and 20% of GMT corresponding to 50% protective antibody titre is shown by the dashed lines, respectively.

**Supplementary Figure 2.**
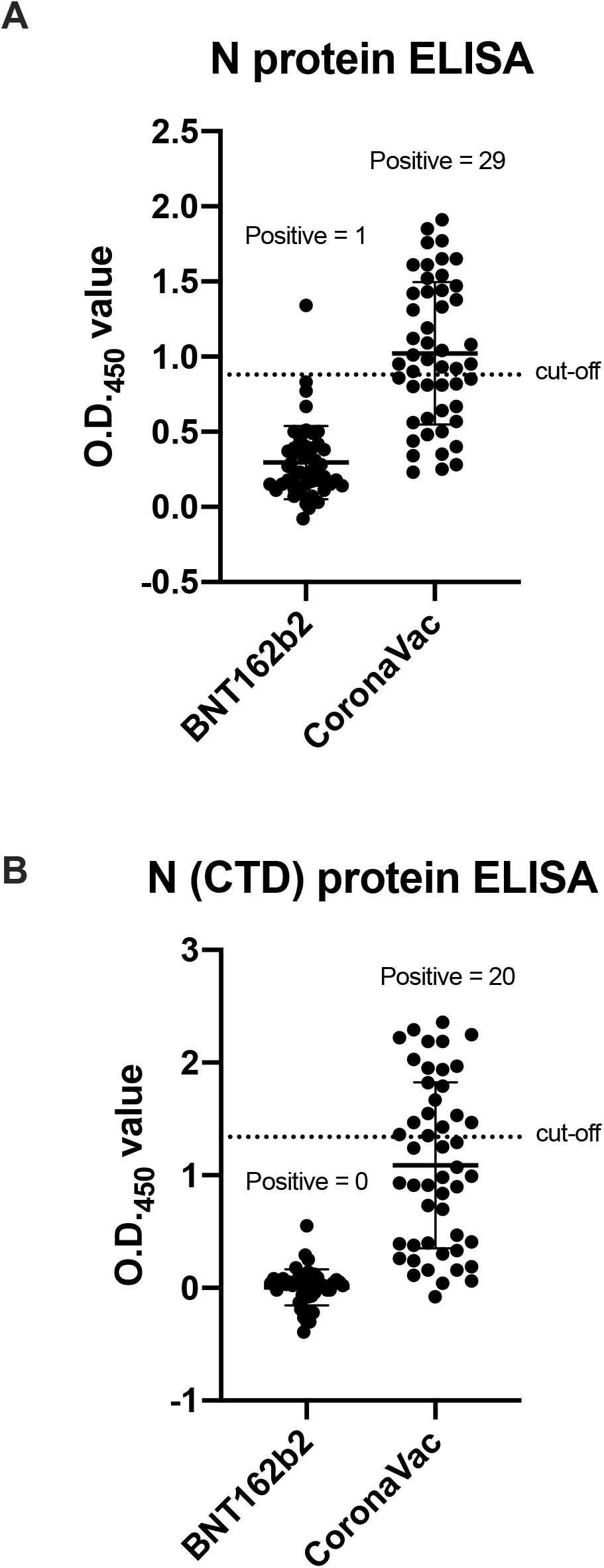
Comparison of antibody responses to SARS-CoV-2 nucleoprotein from the vaccinated cohort. The levels of SARS-CoV-2 (A) full length (N) or (B) C-terminal domain (CTD) of nucleoprotein antibodies were detected from the plasma collected at 1 month after received two doses of BNT162b2 (n=49) or CoronaVac (n=49). Dotted line represents the negative cut-off value.

**Supplementary Figure 3.**
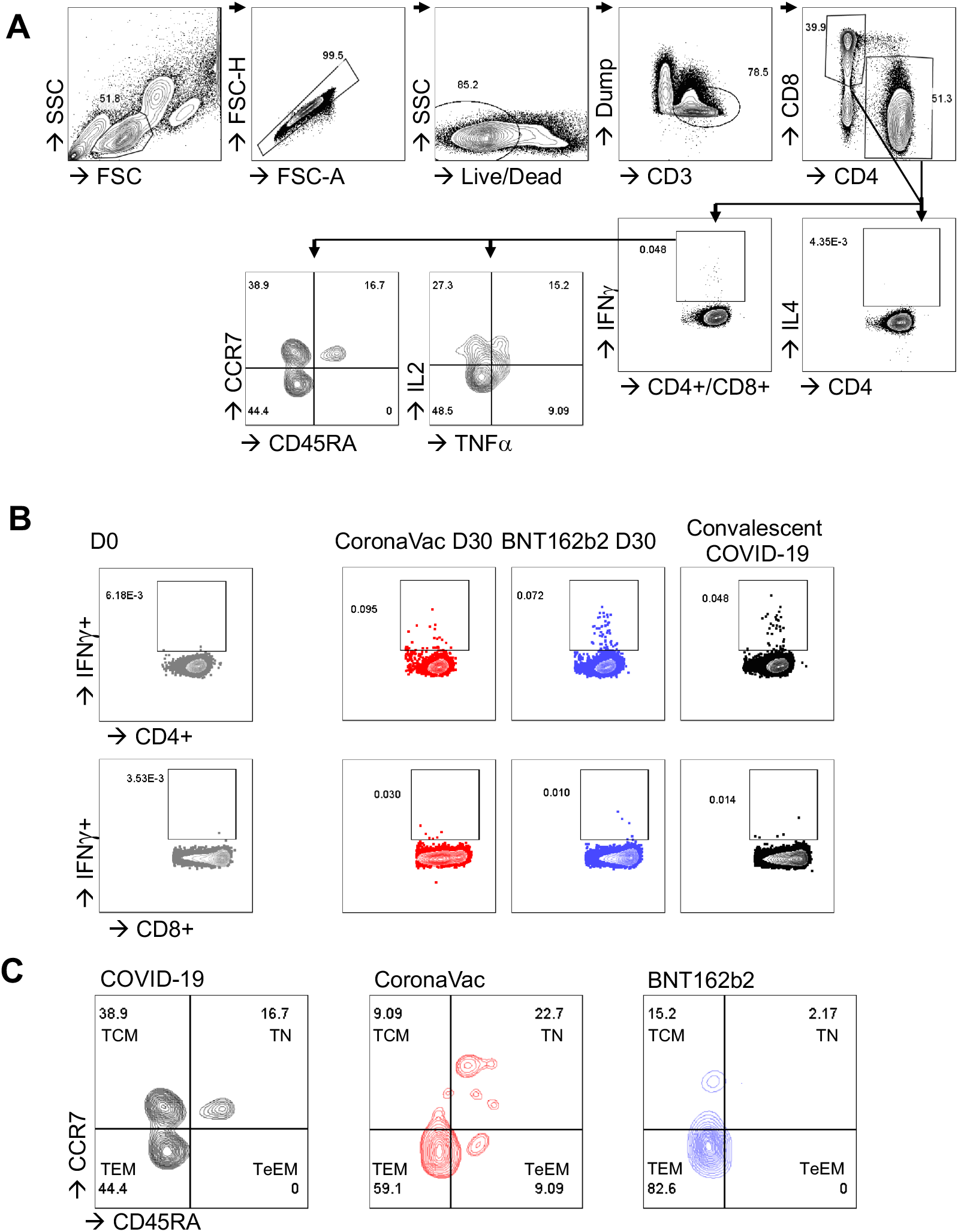
SARS-CoV-2 Spike specific T cells by Flow Cytometry. (A) Gating strategy for characterization of CD4+ and CD8+ T cells by cytokine production (IFNγ, TNFα, IL2, IL4) or memory phenotype (CCR7, CD45RA). (B) Representative FACS plots of IFNg+ responses (B) and memory phenotypes (C) post-SARS-CoV-2 infection or vaccination with inactivated CoronaVac or mRNA vaccine BNT162b2.

**Supplementary Figure 4.**
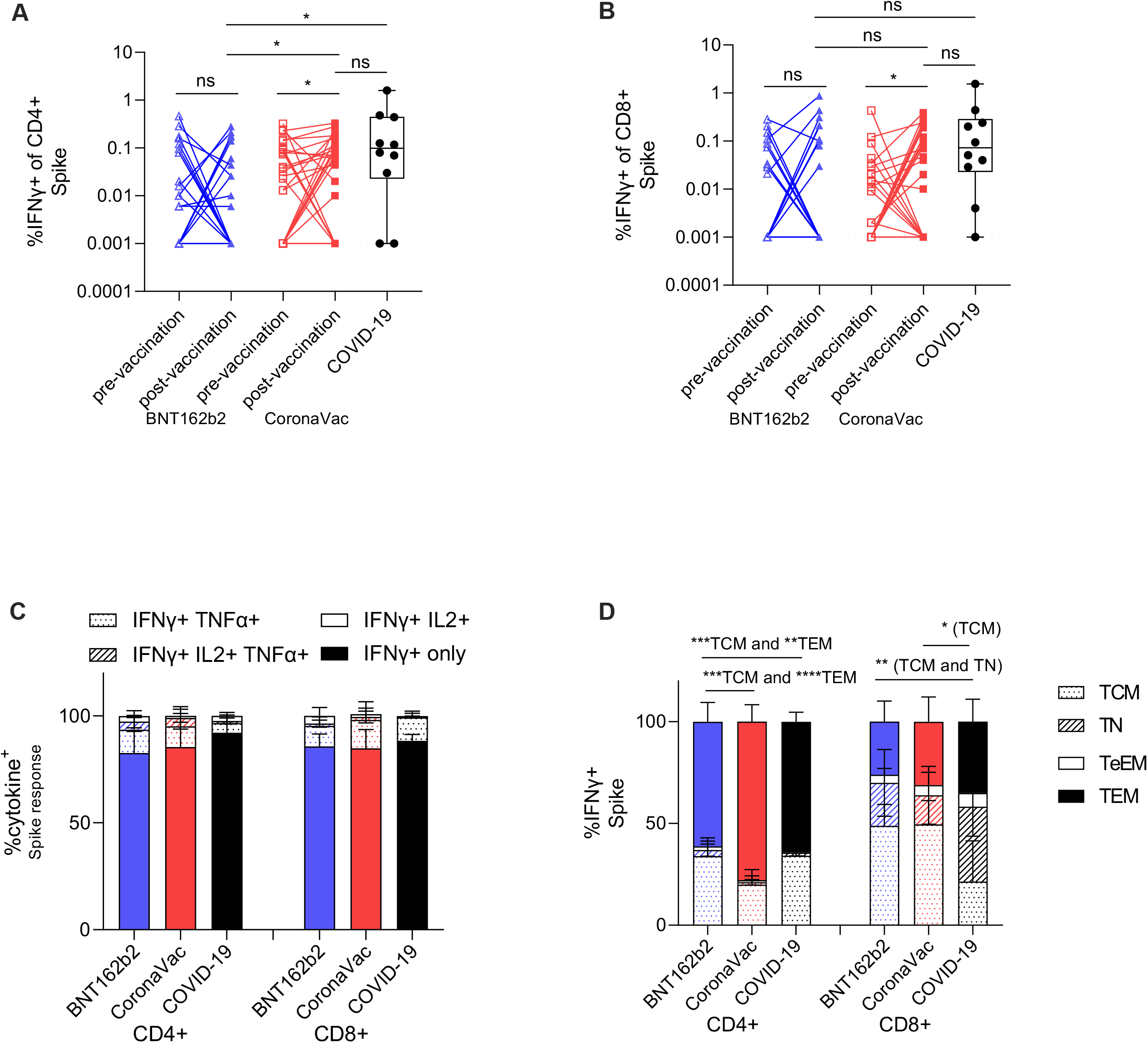
PBMCs from pre- (day 0) and post-vaccination (day 30 post after second dose) of BNT162b2 mRNA (pre-vaccine n=25, post-vaccine n=25) and CoronaVac (pre-vaccine n=30, post-vaccine n=30) and recovered COVID-19 cases (n=10, timepoint 59 days post infection +/- 20 days (mean+/- SD)) were stimulated with S peptides or a DMSO control. The percentage of (A) IFNγ^+^CD4^+^ and (B) IFNγ^+^CD8^+^ T cell were measured by flow cytometry. Dotted lines represent limit of detection following DMSO background subtraction (IFNγ of CD4^+^=0.001, IFNγ of CD8^+^=0.001. (C) The proportion of IFNγ producing IL2 and TNFα CD4^+^ and CD8^+^ T cells post vaccination. (D) The phenotype (by CCR7 and CD45RA) of IFNγ responses for T effector memory (TEM), central memory (TCM), terminal effector memory (TeEM) or naïve (TN) CD4^+^ and CD8^+^ T cells post vaccination. Bars represent the mean values, and error bars represent SD. Statistical significance was determined by paired t-test between pre-versus post-vaccination timepoint samples, and Kruskal-Wallis test for multiple comparisons between vaccines, and COVID-19 patients. *p<0.05, **<0.01, ***<0.001.

**Supplementary Table 1.**
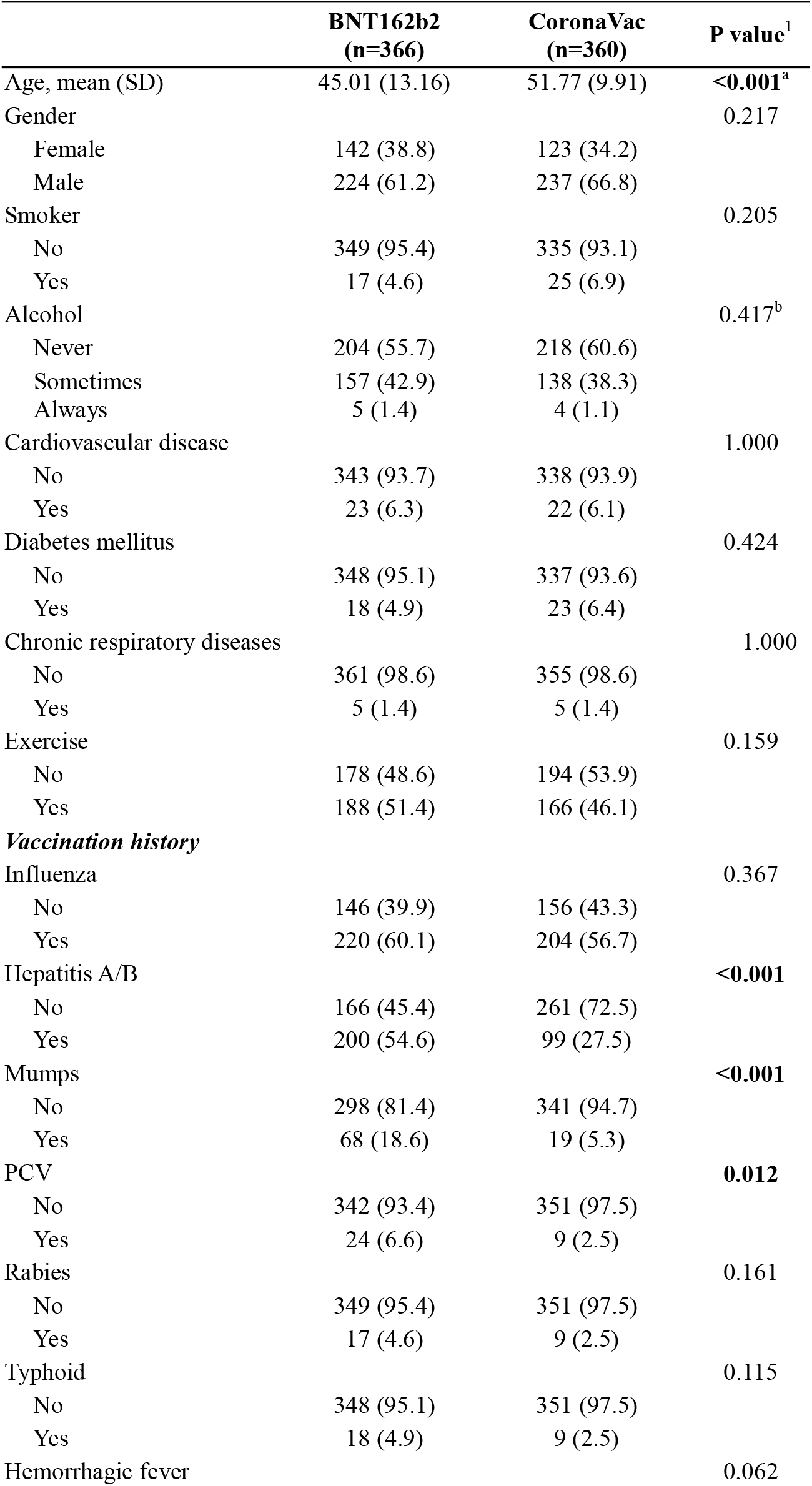
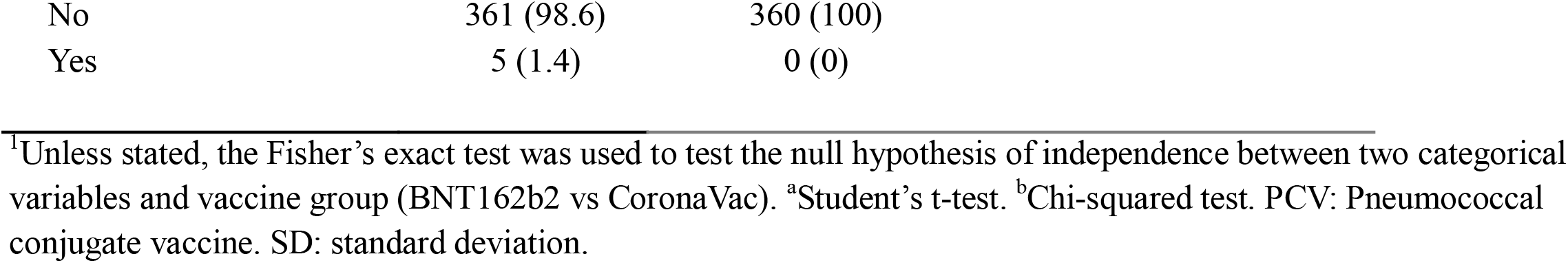
Comparison of characteristics between the two vaccine groups (n=726)..

|  | **BNT162b2**<br>**(n=366)** | **CoronaVac**<br>**(n=360)** | **P value<sup>1</sup>** |
| --- | --- | --- | --- |
| Age, mean (SD) | 45.01 (13.16) | 51.77 (9.91) | **<0.001<sup>a</sup>** |
| Gender |  |  | 0.217 |
| Female | 142 (38.8) | 123 (34.2) |  |
| Male | 224 (61.2) | 237 (66.8) |  |
| Smoker |  |  | 0.205 |
| No | 349 (95.4) | 335 (93.1) |  |
| Yes | 17 (4.6) | 25 (6.9) |  |
| Alcohol |  |  | 0.417<sup>b</sup> |
| Never | 204 (55.7) | 218 (60.6) |  |
| Sometimes | 157 (42.9) | 138 (38.3) |  |
| Always | 5 (1.4) | 4 (1.1) |  |
| Cardiovascular disease |  |  | 1.000 |
| No | 343 (93.7) | 338 (93.9) |  |
| Yes | 23 (6.3) | 22 (6.1) |  |
| Diabetes mellitus |  |  | 0.424 |
| No | 348 (95.1) | 337 (93.6) |  |
| Yes | 18 (4.9) | 23 (6.4) |  |
| Chronic respiratory diseases |  |  | 1.000 |
| No | 361 (98.6) | 355 (98.6) |  |
| Yes | 5 (1.4) | 5 (1.4) |  |
| Exercise |  |  | 0.159 |
| No | 178 (48.6) | 194 (53.9) |  |
| Yes | 188 (51.4) | 166 (46.1) |  |
| ***Vaccination history*** |  |  |  |
| Influenza |  |  | 0.367 |
| No | 146 (39.9) | 156 (43.3) |  |
| Yes | 220 (60.1) | 204 (56.7) |  |
| Hepatitis A/B |  |  | **<0.001** |
| No | 166 (45.4) | 261 (72.5) |  |
| Yes | 200 (54.6) | 99 (27.5) |  |
| Mumps |  |  | **<0.001** |
| No | 298 (81.4) | 341 (94.7) |  |
| Yes | 68 (18.6) | 19 (5.3) |  |
| PCV |  |  | **0.012** |
| No | 342 (93.4) | 351 (97.5) |  |
| Yes | 24 (6.6) | 9 (2.5) |  |
| Rabies |  |  | 0.161 |
| No | 349 (95.4) | 351 (97.5) |  |
| Yes | 17 (4.6) | 9 (2.5) |  |
| Typhoid |  |  | 0.115 |
| No | 348 (95.1) | 351 (97.5) |  |
| Yes | 18 (4.9) | 9 (2.5) |  |
| Hemorrhagic fever |  |  | 0.062 |

|  |  |  |
| --- | --- | --- |
| No | 361 (98.6) | 360 (100) |
| Yes | 5 (1.4) | 0 (0) |

<sup>†</sup>Unless stated, the Fisher's exact test was used to test the null hypothesis of independence between two categorical variables and vaccine group (BNT162b2 vs CoronaVac). <sup>a</sup>Student's t-test. <sup>b</sup>Chi-squared test. PCV: Pneumococcal conjugate vaccine. SD: standard deviation.

**Supplementary Table 2.** Multivariate analysis of factors associated with PRNT_90_ (n=97)

|  | Crude coeff. (95% CI) | Adj. coeff. (95% CI) | P-value |
| --- | --- | --- | --- |
| Vaccine: CoronaVac vs BNT162b2 | -97.4 (-125.0, -69.8) | -95.3 (-123.7, -66.9) | < 0.001 |
| Age | -1.92 (-3.99, 0.16) | -2.36 (-4.14, -0.57) | 0.01 |
| Gender: Male vs Female | 17.9 (-15.8, 51.7) | 7.32 (-21.2, 35.8) | 0.611 |
| Influenza vaccination: Yes vs No | -28.3 (-69.9, 13.3) | -9.74 (-45.6, 26.1) | 0.591 |

One record was excluded from multivariate regression analysis due to incomplete data

**Supplementary Table 3.** Factors associated with PRNT_90_ among subjects given the same vaccine (multivariate)

| Term (BNT162b2) | Adj. coeff. (95% CI) | P-value | Term (CoronaVac) | Adj. coeff. (95% CI) | P-value |
| --- | --- | --- | --- | --- | --- |
| Age (years) | -3.51 (-6.82, -0.19) | 0.039 | Age (years) | -1.48 (-2.92, -0.05) | 0.043 |
| Gender: Male vs Female | 31.7 (-23.7, 87.2) | 0.255 | Gender: Male vs Female | -8.05 (-31.2, 15.1) | 0.487 |

## Notes

**Funding source:** This research was supported by grants from the Health and Medical Research Fund Commissioned Research on the Novel Coronavirus Disease (COVID-19), Hong Kong SAR (COVID1903003) (CKPM, SSN, GCL, MP and DSH), (COVID-190115 and COVID-190126, SAV), Guangdong Province International Scientific and Technological Cooperation Projects (2020A0505100063) (CKPM), the National Research Foundation of Korea (NRF) grant funded through the Korea government (NRF-2018M3A9H4055203) (CKPM), US National Institutes of Health (contract no. HHSN272201400006C) (MP), National Natural Science Foundation of China (NSFC)/Research Grants Council (RGC) Joint Research Scheme (N_HKU737/18) (CKPM and MP) and RGC theme-based research scheme (T11-712/19-N and T11-705/21-N) (DSH). Research was partly supported by Fast Grant ##2161 (Emergent Ventures) to G.K.A., and NIH grants (P01AI120943, R01AI123926 to G.K.A.; R01AI107056 to D.W.L.).

**Conflict of Interest** We declare no competing interests.

### Competing Interest Statement

The authors have declared no competing interest.

### Funding Statement

This research was supported by grants from the Health and Medical Research Fund Commissioned Research on the Novel Coronavirus Disease (COVID-19), Hong Kong SAR (COVID1903003) (CKPM, SSN, GCL, MP and DSH), (COVID-190115 and COVID-190126, SAV), Guangdong Province International Scientific and Technological Cooperation Projects (2020A0505100063) (CKPM), the National Research Foundation of Korea (NRF) grant funded through the Korea government (NRF-2018M3A9H4055203) (CKPM), US National Institutes of Health (contract no. HHSN272201400006C) (MP), National Natural Science Foundation of China (NSFC)/Research Grants Council (RGC) Joint Research Scheme (N_HKU737/18) (CKPM and MP) and RGC theme-based research scheme (T11-712/19-N and T11-705/21-N) (DSH). Research was partly supported by Fast Grant ##2161 (Emergent Ventures) to G.K.A., and NIH grants (P01AI120943, R01AI123926 to G.K.A.; R01AI107056 to D.W.L.).

### Author Declarations

The study was approved by the Joint Chinese University of Hong Kong-New Territories East Cluster Clinical Research Ethics Committee (Ref no: 2020.229) and all participants provided written consent.

## REFERENCES

1) Huang C, Wang Y, Li X, Ren L, Zhao J, Hu Y, Zhang L, Fan G, Xu J, Gu X, Cheng Z, Yu T, Xia J, Wei Y, Wu W, Xie X, Yin W, Li H, Liu M, Xiao Y, Gao H, Guo L, Xie J, Wang G, Jiang R, Gao Z, Jin Q, Wang J, Cao B. (2020) Clinical features of patients infected with 2019 novel coronavirus in Wuhan, China. Lancet. 395(10223):497–506.

2) Guan WJ, Ni ZY, Hu Y, Liang WH, Ou CQ, He JX, Liu L, Shan H, Lei CL, Hui DSC, D. B, Li LJ, Zeng G, Yuen KY, Chen RC, Tang CL, Wang T, Chen PY, Xiang J, Li SY, Wang JL, Liang ZJ, Peng YX, Wei L, Liu Y, Hu YH, Peng P, Wang JM, Liu JY, Chen Z, Li G, Zheng ZJ, Qiu SQ, Luo J, Ye CJ, Zhu SY, Zhong NS; China Medical Treatment Expert Group for Covid-19. (2020) Clinical Characteristics of Coronavirus Disease 2019 in China. N Engl J Med. 382(18):1708–1720.

3) WHO website. https://www.who.int/emergencies/diseases/novel-coronavirus-2019. Access on 25/10/2021.

4) Addetia A, Crawford KHD, Dingens A, Zhu H, Roychoudhury P, Huang ML, Jerome KR, Bloom JD, Greninger AL. (2020) Neutralizing Antibodies Correlate with Protection from SARS-CoV-2 in Humans during a Fishery Vessel Outbreak with a High Attack Rate. J Clin Microbiol. 58(11):e02107–20.

5) Khoury DS, Cromer D, Reynaldi A, Schlub TE, Wheatley AK, Juno JA, Subbarao K, Kent SJ, Triccas JA, Davenport MP. (2021) Neutralizing antibody levels are highly predictive of immune protection from symptomatic SARS-CoV-2 infection. Nat Med. 27(7):1205–1211

6) Corbett KS, Nason MC, Flach B, Gagne M, O’ Connell S, Johnston TS, Shah SN, Edara VV, Floyd K, Lai L, McDanal C, Francica JR, Flynn B, Wu K, Choi A, Koch M, Abiona OM, Werner AP, Alvarado GS, Andrew SF, Donaldson MM, Fintzi J, Flebbe DR, Lamb E, Noe AT, Nurmukhambetova ST, Provost SJ, Cook A, Dodson A, Faudree A, Greenhouse J, Kar S, Pessaint L, Porto M, Steingrebe K, Valentin D, Zouantcha S, Bock KW, Minai M, Nagata BM, Moliva JI, van de Wetering R, Boyoglu-Barnum S, Leung K, Shi W, Yang ES, Zhang Y, Todd JM, Wang L, Andersen H, Foulds KE, Edwards DK, Mascola JR, Moore IN, Lewis MG, Carfi A, Montefiori D, Suthar MS, McDermott A, Sullivan NJ, Roederer M, Douek DC, Graham BS, Seder RA. (2021) Immune Correlates of Protection by mRNA-1273 Immunization against SARS-CoV-2 Infection in Nonhuman Primates. bioRxiv. Preprint.

7) Rodda LB, Netland J, Shehata L, Pruner KB, Morawski PA, Thouvenel CD, Takehara KK, Eggenberger J, Hemann EA, Waterman HR, Fahning ML, Chen Y, Hale M, Rathe J, Stokes C, Wrenn S, Fiala B, Carter L, Hamerman JA, King NP, Gale M Jr, Campbell DJ, Rawlings DJ, Pepper M. (2021) Functional SARS-CoV-2-Specific Immune Memory Persists after Mild COVID-19. Cell. 184(1):169–183.e17.

8) Dan JM, Mateus J, Kato Y, Hastie KM, Yu ED, Faliti CE, Grifoni A, Ramirez SI, Haupt S, Frazier A, Nakao C, Rayaprolu V, Rawlings SA, Peters B, Krammer F, Simon V, Saphire EO, Smith DM, Weiskopf D, Sette A, Crotty S. (2021) Immunological memory to SARS-CoV-2 assessed for up to 8 months after infection. Science. 371(6529):eabf4063.

9) Rydyznski Moderbacher C, Ramirez SI, Dan JM, Grifoni A, Hastie KM, Weiskopf D, Belanger S, Abbott RK, Kim C, Choi J, Kato Y, Crotty EG, Kim C, Rawlings SA, Mateus J, Tse LPV, Frazier A, Baric R, Peters B, Greenbaum J, Ollmann Saphire E, Smith DM, Sette A, Crotty S. (2020) Antigen-Specific Adaptive Immunity to SARS-CoV-2 in Acute COVID-19 and Associations with Age and Disease Severity. Cell. 2183(4):996–1012.e19.

10) Tan AT, Linster M, Tan CW, Le Bert N, Chia WN, Kunasegaran K, Zhuang Y, Tham CYL, Chia A, Smith GJD, Young B, Kalimuddin S, Low JGH, Lye D, Wang LF, Bertoletti A. (2021) Early induction of functional SARS-CoV-2-specific T cells associates with rapid viral clearance and mild disease in COVID-19 patients. Cell Rep. 34(6):108728.

11) Muñoz-Fontela C, Dowling WE, Funnell SGP, Gsell PS, Riveros-Balta AX, Albrecht RA, Andersen H, Baric RS, Carroll MW, Cavaleri M, Qin C, Crozier I, Dallmeier K, de Waal L, de Wit E, Delang L, Dohm E, Duprex WP, Falzarano D, Finch CL, Frieman MB, Graham BS, Gralinski LE, Guilfoyle K, Haagmans BL, Hamilton GA, Hartman AL, Herfst S, Kaptein SJF, Klimstra WB, Knezevic I, Krause PR, Kuhn JH, Le Grand R, Lewis MG, Liu WC, Maisonnasse P, McElroy AK, Munster V, Oreshkova N, Rasmussen AL, Rocha-Pereira J, Rockx B, Rodríguez E, Rogers TF, Salguero FJ, Schotsaert M, Stittelaar KJ, Thibaut HJ, Tseng CT, Vergara-Alert J, Beer M, Brasel T, Chan JFW, García-Sastre A, Neyts J, Perlman S, Reed DS, Richt JA, Roy CJ, Segalés J, Vasan SS, Henao-Restrepo AM, Barouch DH. (2020) Animal models for COVID-19. Nature. 586(7830):509–515.

12) Pormohammad A, Zarei M, Ghorbani S, Mohammadi M, Razizadeh MH, Turner DL, Turner RJ. (2021) Efficacy and Safety of COVID-19 Vaccines: A Systematic Review and Meta-Analysis of Randomized Clinical Trials. Vaccines (Basel). 9(5):467.

13) Walsh EE, Frenck RW Jr, Falsey AR, Kitchin N, Absalon J, Gurtman A, Lockhart S, Neuzil K, Mulligan MJ, Bailey R, Swanson KA, Li P, Koury K, Kalina W, Cooper D, Fontes-Garfias C, Shi PY, Türeci Ö, Tompkins KR, Lyke KE, Raabe V, Dormitzer PR, Jansen KU, Şahin U, Gruber WC. (2020) Safety and Immunogenicity of Two RNA-Based Covid-19 Vaccine Candidates. N Engl J Med. 383(25):2439–2450.

14) Bueno SM, Abarca K, González PA, Gálvez NMS, Soto JA, Duarte LF, Schultz BM, Pacheco GA, González LA, Vázquez Y, Ríos M, Melo-González F, Rivera-Pérez D, Iturriaga C, Urzúa M, Dominguez A, Andrade CA, Berrios RV, Canedo-Marroquín G, Covián C, Moreno-Tapia D, Saavedra F, Vallejos OP, Donato P, Espinoza P, Fuentes D, González M, Guzmán P, Muñoz-Venturelli P, Pérez CM, Potin M, Rojas A, Fasce R, Fernández J, Mora J, Ramírez E, Gaete-Argel A, Oyarzún-Arrau A, Valiente-Echeverría F, Soto-Rifo R, Weiskopf D, Sette A, Zeng G, Meng W, González-Aramundiz JV, Kalergis AM. (2021) Interim report: Safety and immunogenicity of an inactivated vaccine against SARS-CoV-2 in healthy chilean adults in a phase 3 clinical trial. Preprint.

15) Perera RAPM, Ko R, Tsang OTY, Hui DSC, Kwan MYM, Brackman CJ, To EMW, Yen HL, Leung K, Cheng SMS, Chan KH, Chan KCK, Li KC, Saif L, Barrs VR, Wu JT, Sit THC, Poon LLM, Peiris M. (2021) Evaluation of a SARS-CoV-2 Surrogate Virus Neutralization Test for Detection of Antibody in Human, Canine, Cat, and Hamster Sera. J Clin Microbiol. 59(2):e02504–20.

16) Lau EHY, Hui DSC, Tsang OTY, Chan WH, Kwan MYK, Chiu SS, Cheng SMS, Ko RLW, Li JKC, Chaothai S, Tsang CH, Poon LLM, Peiris M. Long-term persistence of SARS-CoV-2 neutralizing antibody responses after infection and estimates of the duration of protection. EClinicalMedicine. In press.

17) Pan H, Wu Q, Zeng G, Yang J, Jiang D, Deng X, Chu K, Zheng W, Zhu F, Yu H, Yin W. Immunogenicity and safety of a third dose, and immune persistence of CoronaVac vaccine in healthy adults aged 18-59 years: interim results from a double-blind, randomized, placebo-controlled phase 2 clinical trial. medRxiv July 25 2021. doi: https://doi.org/10.1101/2021.07.23.21261026

18) Ju B, Zhang Q, Ge J, Wang R, Sun J, Ge X, Yu J, Shan S, Zhou B, Song S, Tang X, Yu J, Lan J, Yuan J, Wang H, Zhao J, Zhang S, Wang Y, Shi X, Liu L, Zhao J, Wang X, Zhang Z, Zhang L. (2020) Human neutralizing antibodies elicited by SARS-CoV-2 infection. Nature. 584(7819):115–119.

19) Chi X, Yan R, Zhang J, Zhang G, Zhang Y, Hao M, Zhang Z, Fan P, Dong Y, Yang Y, Chen Z, Guo Y, Zhang J, Li Y, Song X, Chen Y, Xia L, Fu L, Hou L, Xu J, Yu C, Li J, Zhou Q, Chen W. (2020) A neutralizing human antibody binds to the N-terminal domain of the Spike protein of SARS-CoV-2. Science. 369(6504):650–655.

20) Andreano E, Nicastri E, Paciello I, Pileri P, Manganaro N, Piccini G, Manenti A, Pantano E, Kabanova A, Troisi M, Vacca F, Cardamone D, De Santi C, Torres JL, Ozorowski G, Benincasa L, Jang H, Di Genova C, Depau L, Brunetti J, Agrati C, Capobianchi MR, Castilletti C, Emiliozzi A, Fabbiani M, Montagnani F, Bracci L, Sautto G, Ross TM, Montomoli E, Temperton N, Ward AB, Sala C, Ippolito G, Rappuoli R. (2021) Extremely potent human monoclonal antibodies from COVID-19 convalescent patients. Cell. 184(7):1821–1835.e16.

21) McLean MR, Madhavi V, Wines BD, Hogarth PM, Chung AW, Kent SJ. (2017) Dimeric Fcγ Receptor Enzyme-Linked Immunosorbent Assay To Study HIV-Specific Antibodies: A New Look into Breadth of Fcγ Receptor Antibodies Induced by the RV144 Vaccine Trial. J Immunol. 199(2):816–826.

22) Kaplonek P, Cizmeci D, Fischinger S, Collier AR, Suscovich T, Linde C, Broge T, Mann C, Amanat F, Dayal D, Rhee J, de St Aubin M, Nilles EJ, Musk ER, Menon AS, Saphire EO, Krammer F, Lauffenburger DA, Barouch DH, Alter G. (2021) Subtle immunological differences in mRNA-1273 and BNT162b2 COVID-19 vaccine induced Fc-functional profiles. bioRxiv

23) Gorman MJ, Patel N, Guebre-Xabier M, Zhu AL, Atyeo C, Pullen KM, Loos C, Goez-Gazi Y, Carrion R Jr, Tian JH, Yuan D, Bowman KA, Zhou B, Maciejewski S, McGrath ME, Logue J, Frieman MB, Montefiori D, Mann C, Schendel S, Amanat F, Krammer F, Saphire EO, Lauffenburger DA, Greene AM, Portnoff AD, Massare MJ, Ellingsworth L, Glenn G, Smith G, Alter G. (2021) Fab and Fc contribute to maximal protection against SARS-CoV-2 following NVX-CoV2373 subunit vaccine with Matrix-M vaccination. Cell Rep Med. 2(9):100405.

24) McMahan K, Yu J, Mercado NB, Loos C, Tostanoski LH, Chandrashekar A, Liu J, Peter L, Atyeo C, Zhu A, Bondzie EA, Dagotto G, Gebre MS, Jacob-Dolan C, Li Z, Nampanya F, Patel S, Pessaint L, Van Ry A, Blade K, Yalley-Ogunro J, Cabus M, Brown R, Cook A, Teow E, Andersen H, Lewis MG, Lauffenburger DA, Alter G, Barouch DH. (2021) Correlates of protection against SARS-CoV-2 in rhesus macaques. Nature. 590(7847):630–634.

25) Atyeo C, Fischinger S, Zohar T, Slein MD, Burke J, Loos C, McCulloch DJ, Newman KL, Wolf C, Yu J, Shuey K, Feldman J, Hauser BM, Caradonna T, Schmidt AG, Suscovich TJ, Linde C, Cai Y, Barouch D, Ryan ET, Charles RC, Lauffenburger D, Chu H, Alter G. (2020) Distinct Early Serological Signatures Track with SARS-CoV-2 Survival. Immunity. 53(3):524–532.e4.

26) Lu J, Mold C, Du Clos TW, Sun PD. (2018) Pentraxins and Fc Receptor-Mediated Immune Responses. Front Immunol. 9:2607.

27) Polack FP, Thomas SJ, Kitchin N, Absalon J, Gurtman A, Lockhart S, Perez JL, Pérez Marc G, Moreira ED, Zerbini C, Bailey R, Swanson KA, Roychoudhury S, Koury K, Li P, Kalina WV, Cooper D, Frenck RW Jr, Hammitt LL, Türeci Ö, Nell H, Schaefer A, Ünal S, Tresnan DB, Mather S, Dormitzer PR, Şahin U, Jansen KU, Gruber WC; C4591001 Clinical Trial Group. (2020) Safety and Efficacy of the BNT162b2 mRNA Covid-19 Vaccine. N Engl J Med. 383(27):2603–2615.

28) Haas EJ, Angulo FJ, McLaughlin JM, Anis E, Singer SR, Khan F, Brooks N, Smaja M, Mircus G, Pan K, Southern J, Swerdlow DL, Jodar L, Levy Y, Alroy-Preis S. (2021) Impact and effectiveness of mRNA BNT162b2 vaccine against SARS-CoV-2 infections and COVID-19 cases, hospitalisations, and deaths following a nationwide vaccination campaign in Israel: an observational study using national surveillance data. Lancet. 397(10287):1819–1829.

29) Recommendation for an emergency use listing of CODI-19 vaccine (Vero Cell), inactivated. Submitted by Sinovac. https://extranet.who.int/pqweb/sites/default/files/documents/SINOVAC_TAG_PEG_REPORT_EUL-Final28june2021.pdf

30) Tanriover MD, Doğanay HL, Akova M, Güner HR, Azap A, Akhan S, Köse Ş, Erdinç FŞ, Akalın EH, Tabak ÖF, Pullukçu H, Batum Ö, Şimşek Yavuz S, Turhan Ö, Yıldırmak MT, Köksal İ, Taşova Y, Korten V, Yılmaz G, Çelen MK, Altın S, Çelik İ, Bayındır Y, Karaoğlan İ, Yılmaz A, Özkul A, Gür H, Unal S; CoronaVac Study Group. Efficacy and safety of an inactivated whole-virion SARS-CoV-2 vaccine (CoronaVac): interim results of a double-blind, randomised, placebo-controlled, phase 3 trial in Turkey. (2021) Lancet. 398:213–222.

31) Jara A, Undurraga EA, González C, Paredes F, Fontecilla T, Jara G, Pizarro A, Acevedo J, Leo K, Leon F, Sans C, Leighton P, Suárez P, García-Escorza H, Araos R. (2021) Effectiveness of an Inactivated SARS-CoV-2 Vaccine in Chile. N Engl J Med. In press.

32) McMahan K, Yu J, Mercado NB, Loos C, Tostanoski LH, Chandrashekar A, Liu J, Peter L, Atyeo C, Zhu A, Bondzie EA, Dagotto G, Gebre MS, Jacob-Dolan C, Li Z, Nampanya F, Patel S, Pessaint L, Van Ry A, Blade K, Yalley-Ogunro J, Cabus M, Brown R, Cook A, Teow E, Andersen H, Lewis MG, Lauffenburger DA, Alter G, Barouch DH. (2021) Correlates of protection against SARS-CoV-2 in rhesus macaques. Nature. 590(7847):630–634.

32) Geers D, Shamier MC, Bogers S, den Hartog G, Gommers L, Nieuwkoop NN, Schmitz KS, Rijsbergen LC, van Osch JAT, Dijkhuizen E, Smits G, Comvalius A, van Mourik D, Caniels TG, van Gils MJ, Sanders RW, Oude Munnink BB, Molenkamp R, de Jager HJ, Haagmans BL, de Swart RL, Koopmans MPG, van Binnendijk RS, de Vries RD, GeurtsvanKessel CH. (2021) SARS-CoV-2 variants of concern partially escape humoral but not T-cell responses in COVID-19 convalescent donors and vaccinees. Sci Immunol. 6:eabj1750.

33) Nguyen THO, Rowntree LC, Petersen J, Chua BY, Hensen L, Kedzierski L, van de Sandt CE, Chaurasia P, Tan HX, Habel JR, Zhang W, Allen LF, Earnest L, Mak KY, Juno JA, Wragg K, Mordant FL, Amanat F, Krammer F, Mifsud NA, Doolan DL, Flanagan KL, Sonda S, Kaur J, Wakim LM, Westall GP, James F, Mouhtouris E, Gordon CL, Holmes NE, Smibert OC, Trubiano JA, Cheng AC, Harcourt P, Clifton P, Crawford JC, Thomas PG, Wheatley AK, Kent SJ, Rossjohn J, Torresi J, Kedzierska K. (2021) CD8+ T cells specific for an immunodominant SARS-CoV-2 nucleocapsid epitope display high naive precursor frequency and TCR promiscuity. Immunity. 54:1066–1082.e5.

34) Roberts AD, Ely KH, Woodland DL. (2005) Differential contributions of central and effector memory T cells to recall responses. J Exp Med. 202:123–33.

35) Wrapp D, Wang N, Corbett KS, Goldsmith JA, Hsieh CL, Abiona O, Graham BS, McLellan JS. (2020) Cryo-EM structure of the 2019-nCoV spike in the prefusion conformation. Science. 367(6483):1260–1263.

36) Zhang Y, Zeng G, Pan H, Li C, Hu Y, Chu K, Han W, Chen Z, Tang R, Yin W, Chen X, Hu Y, Liu X, Jiang C, Li J, Yang M, Song Y, Wang X, Gao Q, Zhu F. (2021) Safety, tolerability, and immunogenicity of an inactivated SARS-CoV-2 vaccine in healthy adults aged 18-59 years: a randomised, double-blind, placebo-controlled, phase 1/2 clinical trial. Lancet Infect Dis. 21:181–192.

37) Lim WW, Mak L, Leung GM, Cowling BJ, Peiris M. Comparative immunogenicity of mRNA and inactivated vaccines against COVID-19. Lancet Microbe. Epub ahead of print.

