## Supplementary file for "Comparison of the immunogenicity of BNT162b2 and CoronaVac COVID-19 Vaccines in Hong Kong"

Online Supplemental file

**Comparison of the immunogenicity of BNT162b2 and CoronaVac COVID-19 Vaccines in Hong Kong: An observational cohort study.**

Chris Ka Pun Mok^1,2,*^, Carolyn A Cohen^3,*^, Samuel M.S. Cheng^4,*^, Chunke Chen^1,2^, Kin-On Kwok^1^, Karen Yiu^5^, Tat-On Chan^5^, Maireid Bull^3^, Kwun Cheung Ling^5^, Zixi Dai^3^, Susanna S Ng^5^, Grace Chung-Yan Lui^5,6^, Chao Wu^7^, Gaya K. Amerasinghe^7^, Daisy W Leung^8^, Samuel Yeung Shan Wong^1^, Sophie A Valkenburg^3^, Malik Peiris^3,4,#^, David S Hui^5,6,#^

^1^The Jockey Club School of Public Health and Primary Care, The Chinese University of Hong Kong, Hong Kong SAR, PR China.

^2^Li Ka Shing Institute of Health Sciences, Faculty of Medicine, The Chinese University of Hong Kong, Hong Kong SAR, PR China.

^3^HKU-Pasteur Research Pole, School of Public Health, Li Ka Shing Faculty of Medicine, The University of Hong Kong, Hong Kong SAR, China.

^4^School of Public Health, Li Ka Shing Faculty of Medicine, The University of Hong Kong, Hong Kong SAR, China.

^5^Department of Medicine and Therapeutics, The Chinese University of Hong Kong, Hong Kong SAR, PR China.

^6^Stanley Ho Centre for Emerging Infectious Diseases, Faculty of Medicine, The Chinese University of Hong Kong, Hong Kong SAR, China.

^7^Department of Pathology and Immunology, Washington University School of Medicine in St. Louis, St. Louis, MO, USA

^8^Department of Internal Medicine, Washington University School of Medicine in St. Louis, St. Louis, MO, USA

*equal contribution

#Corresponding authors:

**Malik Peiris,** School of Public Health, Li Ka Shing Faculty of Medicine, The University of Hong Kong, 7 Sassoon Rd, Pokfulam, Hong Kong Special Administrative Region, Republic of China, Tel: +852-39177537,

**David S Hui,** Department of Medicine and Therapeutics, The Chinese University of Hong Kong, Prince of Wales Hospital, Hong Kong Special Administrative Region, Republic of China, Tel: +852-35053128,

**Processing and storage of specimens.** The blood samples were first centrifuged at 3000 xg for 10 minutes at room temperature for plasma collection. The remaining blood was diluted with equal volume of PBS buffer, transferred onto the Ficoll-Paque Plus medium (GE Healthcare), and centrifuged at 400 xg for 20 minutes. Peripheral Blood Mononuclear Cells (PBMC) samples were then collected and washed with cold RPMI-1640 medium. The plasma and PBMC were stored at -80°C and liquid nitrogen respectively until use. Plasma samples were heat inactivated (HI) for 30 minutes at 56^o^C prior to use.

**Protein expression and purification.** The receptor-binding domain (RBD, residues 319-541) and N-terminal domain (NTD, residues 14-305) of the SARS-CoV-2 spike protein (GenBank: QHD43416.1) were cloned into a customized pFastBac vector. The RBD and NTD constructs were fused with an N-terminal gp67 signal peptide and a C-terminal His6 tag. Recombinant bacmid DNA was generated using the Bac-to-Bac system (Life Technologies, Thermo Fisher Scientific). Baculovirus was generated by transfecting purified bacmid DNA into Sf9 cells using FuGENE HD (Promega, Madison, US) and subsequently used to infect suspension cultures of High Five cells (Life Technologies) at a multiplicity of infection (MOI) of 5 to 10. Infected High Five cells were incubated at 28 °C with shaking at 110 rpm for 72 hours for protein expression. The supernatant was then concentrated using a Centramate cassette (10 kDa molecular weight cutoff for RBD, Pall Corporation, New York, USA). RBD and NTD proteins were purified by Ni-NTA Superflow (Qiagen, Hilden, Germany), followed by size exclusion chromatography and buffer exchange to phosphate-buffered saline (PBS). The S2 protein were purchased from Sino Biological, China.

SARS-CoV-2 N constructs were expressed as His-tag fusion proteins in BL21 (DE3) E. coli cells (Novagen). At OD_600_ of 0.6-0.7, recombinant protein expression was induced with 0.5 mM isopropyl β-d-1-thiogalactopyranoside (IPTG) for 12-14 hours at 18˚C. Cells were harvested and resuspended in lysis buffer containing 20 mM Tris (pH 7.5), 1 M NaCl, 20 mM imidazole, 5 mM 2-mecaptoethanol (BME). Cells were lysed using an EmulsiFlex-C5 homogenizer (Avestin) and lysates were clarified by centrifugation at 30,000 x g at 4 ˚C for 40 min. N proteins were purified using affinity tag and gel filtration columns. Fractions were pooled and concentrated, then aliquoted and flash frozen in liquid nitrogen. N (CTD, a.a. 248-369) has limited RNA binding, therefore high salt purification condition resulted in RNA-free protein. For N (NTD-LKR-CTD-Carm, a.a. 44-419), because of high affinity RNA binding even at high salt condition. both RNA-bound and RNA-free states were purified, and only RNA-free fractions (A_260/280_ ~ 0.5) were used for ELISA studies. Purity of N proteins were determined by Coomassie staining of SDS-PAGE.

**ELISA for spike RBD, NTD, S2 and N protein antibodies.** A 96-well enzyme-linked immunosorbent assay (ELISA) plate (Nunc MaxiSorp, Thermo Fisher Scientific) was first coated overnight with 100 ng per well of purified recombinant protein in PBS buffer. The plates were then blocked with 100 μl of Chonblock blocking/sample dilution ELISA buffer (Chondrex Inc, Redmon, US) and incubated at room temperature for 1 hour. Each human plasma sample was diluted to 1:100 in Chonblock blocking/sample dilution ELISA buffer. Each sample was then added into the ELISA plates for a two-hour incubation at 37°C. After extensive washing with PBS containing 0.1% Tween 20, each well in the plate was further incubated with the anti-human IgG secondary antibody (1:5000, Thermo Fisher Scientific) for 1 hour at 37°C. The ELISA plates were then washed five times with PBS containing 0.1% Tween 20. Subsequently, 100 μl of HRP substrate (Ncm TMB One; New Cell and Molecular Biotech Co. Ltd, Suzhou, China) was added into each well. After 15 min of incubation, the reaction was stopped by adding 50 μl of 2 M H_2_SO_4_ solution and analyzed on an absorbance microplate reader at 450 nm wavelength (1).

**ELISA for FcγRIIIa and Avidity**

Plates (Nunc MaxiSorp, Thermofisher Scientific) were coated with 80 ng/ml S or N protein (SinoBiological). Plates were rinsed, blocked with 1% FBS in PBS, incubated with 1:100 HI plasma diluted in 0.05% Tween-20/ 0.1% FBS in PBS for 2 hours then rinsed again. To measure assess antibody avidity binding, an additional 3 washes with 8M Urea was included before incubation of samples for 2 hours with IgG-HRP (1:5000, G8-185; BD). To measure antibody FcγRIIIa-binding, plates were instead coated with proteins at 500 ng/ml, followed by HI plasma at 1:50. Instead of IgG-HRP, plates were incubated with biotinylated dimeric FcgRIIIa-V158 (2) at 50 ng/ml for 1 hour at 37°C, rinsed and incubated with Streptavidin-HRP (1:10000, Pierce). HRP was revealed by stabilized hydrogen peroxide and tetramethylbenzidine (R&D systems) for 20 minutes, stopped with 2 M H_2_SO_4_ and analyzed on an absorbance microplate reader at 450 nm wavelength (Tecan Life Sciences).

**Surrogate virus neutralization test (sVNT).** SARS-CoV-2 surrogate virus neutralization test kits were obtained from GenScript, Inc., NJ, USA, and the tests were carried out according to the manufacturer’s instructions. The test sera (10 μl) and positive and negative controls were diluted 1:10 and mixed with an equal volume of horseradish peroxidase (HRP) conjugated to SARS-CoV-2 spike receptor binding domain (RBD) (6 ng) and incubated for 30 min at 37°C. A 100 μl volume of each mixture was added to each well on the microtiter plate coated with ACE-2 receptor. The plate was sealed and incubated at room temperature for 15 min at 37°C. Plates were then washed with wash solution and tapped dry, and 100 μl of 3,3’,5,5’-tetramethylbenzidine (TMB) solution was added to each well and incubated in the dark at room temperature for 15 min. The reaction was stopped by addition of 50 μl of Stop Solution to each well and the absorbance read at 450 nm in an ELISA microplate reader. The assay validity was based on values representing optical density at 450 nm (OD450) for positive and negative results falling within the range of recommended values. On the basis of the assumption that the positive and negative controls gave the recommended OD_450_ values, percent inhibition of each serum was calculated as follows: percent inhibition (1 - sample OD value/negative-control OD value) x 100. Percent inhibition values of 20% or more are regarded as positive results (3).

**Plaque reduction neutralization test (PRNT).** Plasma samples were two-fold diluted starting from a 1:10 dilution and mixed with equal volumes of around 120 plaque-forming units (pfu) of SARS-CoV-2 as determined by Vero E6. After 1-hour incubation at 37°C, the plasma-virus mixture was added onto cell monolayers seated in a 24-well cell culture plate and incubated for 1 hour at 37°C with 5% CO_2_. The plasma-virus mixtures were then discarded and infected cells were immediately covered with 1% agarose gel in DMEM medium. After incubation for 3 days at 37°C with 5% CO_2_, the plates were formalin fixed and stained by 0.5% crystal violet solution. Neutralization titers were determined by the highest plasma dilution that resulted in >50% (PRNT_50_) or >90% (PRNT_90_) reduction in the number of pfu. These tests were performed in a BSL3 facility at the University of Hong Kong (4).

**SARS-CoV-2-specific T cells by Intracellular Cytokine Staining (ICS).** Cryopreserved PBMCs were thawed and re-stimulated with an overlapping peptide pool representing the SARS-CoV-2 spike proteins (300 nM), structural proteins (spike, nucleocapsid, envelope and membrane) (300nM) or DMSO (1% in RPMI) in two independent experiments. Experiments involving both the structural and spike pools involved cell stimulation for 28 hours at 37^o^C. Golgi Plug (BD) containing Brefeldin A (1% in PBS), and Golgi Stop (BD) containing Monensin (0.67% in PBS) was added at 24 hours during stimulation. A supplementary experiment using only the spike peptide pool involved a shorter cell stimulation for 6 hours at 37^o^C, with Golgi Plug and Golgi Stop (added at 2 hours during stimulation). The amino acid sequence of the peptide pools was based on βCoV/Hong Kong/VM20001061/2020 strain (GISAID ID: EPI_ISL_412028). Cells were stained with Zombie-NIR (all antibodies from Biolegend and clone used) followed by anti-human CD3-PE/Dazzle 594 (UCHT1), CD4-BV605 (OKT4), CD8-AlexaFluor700 (SK1), CCR7-PerCP/Cy5.5 (G043H7), PD-1-BV421 (NAT105), CD45RA-APC (HI100), and a dump channel containing CD19-BV510 (HIB19), CD56-BV510 (5.1H11) and CD14-BV510 (M5E2). Following cell permeabilization, intracellular staining with anti-IFNγ-FITC (4S.B3), IL4-PE (MP4-25D2) and TNF-BV711 (MAb11), IL2-PECy7 (MQ1-17H12), was carried out before acquisition of samples. Stained cells were acquired via flow cytometry (AttuneNxT) and analysed by FlowJo v10. Representative FACS plots and gating strategy are shown in Supplementary Figure 2A and B. Samples were included in subsequent analyses if cell viability was above a 40% cut-off, leading to exclusion of 4 pre-vaccination samples in total (3 from BNT162b2 and 1 from CoronaVac groups).

**Statistical analysis.** Continuous variables were summarized as mean with standard deviation (SD) while categorical variables summarized as frequency with percentage. Geometric means were used for comparison of antibody titers. Comparison between groups was conducted with the rank sum test for continuous variables and the Chi-square test or Fisher’s exact test for categorical variables as appropriate. We employed a multivariate regression model controlled for potential confounders (age and gender) to determine factors associated with PRNT titers. The final model was determined with a stepwise variable selection technique based on the model with the lowest Akaike’s Information Criterion (AIC) value, and retaining only variables with a p-value <0.1. All analyses were performed in R (version 4.1.0; R Foundation for Statistical Computing). Statistical analysis on serology and T cell data was performed on Prism 9 (Graphpad). For two-way comparison, the student paired t-test (paired) or Mann-Whitney t-test (unpaired) was used. For multiple-group comparisons, a Friedman (paired) or Kruskal-Wallis (unpaired) test, followed by the Dunn-Bonferroni post-hoc test was used. P values < 0.05 were considered statistically significant.

**References:**

1) Perera RA, Mok CK, Tsang OT, Lv H, Ko RL, Wu NC, Yuan M, Leung WS, Chan JM, Chik TS, Choi CY, Leung K, Chan KH, Chan KC, Li KC, Wu JT, Wilson IA, Monto AS, Poon LL, Peiris M. (2020) Serological assays for severe acute respiratory syndrome coronavirus 2 (SARS-CoV-2), March 2020. Euro Surveill. 25(16):2000421.

2) Wines BD, Vanderven HA, Esparon SE, Kristensen AB, Kent SJ, Hogarth PM. Dimeric FcγR Ectodomains as Probes of the Fc Receptor Function of Anti-Influenza Virus IgG. J Immunol. 2016 Aug 15;197(4):1507-16. doi: 10.4049/jimmunol.1502551. Epub 2016 Jul 6. PMID: 27385782.

3) Perera RAPM, Ko R, Tsang OTY, Hui DSC, Kwan MYM, Brackman CJ, To EMW, Yen HL, Leung K, Cheng SMS, Chan KH, Chan KCK, Li KC, Saif L, Barrs VR, Wu JT, Sit THC, Poon LLM, Peiris M. (2021) Evaluation of a SARS-CoV-2 Surrogate Virus Neutralization Test for Detection of Antibody in Human, Canine, Cat, and Hamster Sera. J Clin Microbiol. 59(2):e02504-20.

4) Lau EHY, Tsang OTY, Hui DSC, Kwan MYW, Chan WH, Chiu SS, Ko RLW, Chan KH, Cheng SMS, Perera RAPM, Cowling BJ, Poon LLM, Peiris M. (2021) Neutralizing antibody titres in SARS-CoV-2 infections. Nat Commun. 12(1):63.
